# Metabolic Phenotyping Objectively Captures Dietary Intake and Short-term Cardiovascular Disease Risk Responses Under an Inpatient Randomized Crossover Clinical Trial

**DOI:** 10.64898/2026.03.20.26348884

**Authors:** Yiwei Wu, Lina Alqarni, Joram M Posma, Melpomeni Kasapi, Liam Walsh, Orla O’Sullivan, Elaine Holmes, Gary Frost, Isabel Garcia-Perez

## Abstract

**Background:** Diet is central to cardiovascular disease (CVD) prevention, yet free-living studies rarely capture what people eat at home or how closely they follow an assigned diet due to limitations in self-reporting. Short-term inpatient feeding studies, with all meals provided and supervised intake, allow direct assessment of the physiological effects of dietary patterns.

**Objectives:** i) To compare the short-term effects of a UK-National Institute for Health and Care Excellence (NICE) aligned diet versus a Western-style diet on CVD risk factors, metabolic phenotypes and microbiome; ii) To evaluate whether urinary metabolic phenotyping can objectively classify dietary adherence in adults with increased CVD risk.

**Methods:** In a controlled inpatient randomized crossover trial, 18 adults at elevated CVD risk completed two 72 h isocaloric diets: NICE-compliant and Western-style. Repeated-measures MCCV-PLS-DA assessed NMR fasting serum and 24 h urine metabolomic phenotypes. Univariate analyses examined CVD markers, urinary metabolites, serum SCFAs, and gut microbial richness and α-diversity.

**Results:** Diet modulated CVD risk markers, with the NICE compliant diet lowering systolic blood pressure and atherogenic lipid parameters, whereas the Western-style diet increased these measures (all q < 0.05). The Western-style diet reduced microbial richness and tended towards lower α-diversity. Urinary metabolic phenotyping identified 27 discriminatory metabolites between the diets reflecting food intake. Most diet-linked metabolites diverged from baseline within 24 h; microbiome derived metabolites demonstrated early and sustained divergence across 72 h. The urinary MCCV-PLS-DA model extended from a previously published framework in healthy adults, robustly classified dietary adherence (Q^2^Y=0.96), and correctly predicted allocated dietary intervention at earlier timepoints (24-48 h).

**Conclusions:** Urinary metabolic phenotyping offers a sensitive and non-invasive tool for objectively assessing dietary intake. Short-term adherence to contrasting dietary patterns produced rapid, diet-specific metabolic and microbial effect in individuals at elevated CVD risk and differentially impacted cardiovascular risk profiles.

**This trial was registered at the ISRCTN registry (**https://www.isrctn.com/ISRCTN44705179).

## Introduction

Cardiovascular diseases (CVD) remain the leading cause of mortality worldwide, accounting for nearly one-third of all deaths and imposing a substantial burden on healthcare systems globally (1). CVD is preventable through lifestyle modification (2, 3) and adherence to a cardioprotective dietary pattern is consistently associated with lower CVD risk (4, 5). Meta-analyses report that Mediterranean dietary pattern reduces CVD incidence by 10% (4). The Dietary Approaches to Stop Hypertension (DASH) diet lowers blood pressure and cholesterol, predicting a 13% reduction in 10 years CVD risk (5). In the United Kingdom, guidance from the National Institute for Health and Care Excellence (NICE) emphasizes a dietary pattern rich in fruits, vegetables, and whole-grain foods, with reduced intake of saturated fats, salt, and processed foods, as CVD prevention and management. Despite well-established dietary guidelines, the incidence of CVD continues to rise (6). A persistent challenge to CVD prevention is that dietary intake in the home environment is poorly measured. Misreporting (7, 8) using current self-reported dietary methodologies ranges from 30-88%. This makes it difficult to determine whether poor outcomes reflect non-adherence to dietary guidelines or variability in individual response to the guidelines themselves.

Metabolic phenotyping strategies have become a valuable approach to objectively characterize dietary intake and its metabolic effects. High-throughput analytical platforms enable comprehensive quantification of low-molecular-weight metabolites that reflect the intake of specific nutrients, foods, and food groups, complementing self-reported dietary assessment methods (9–12). Additionally, these metabolic profiles capture downstream processing of dietary compounds, enabling evaluation of inter-individual variability arising from genetic, microbial, and other host-related factors (13–14). We previously developed a urinary metabolic phenotyping framework using proton nuclear magnetic resonance (^1^H-NMR) spectroscopy to objectively assess dietary patterns (15). Under tightly controlled inpatient feeding conditions, urinary metabolite signatures discriminated diets differing in adherence to World Health Organization (WHO) healthy eating guidelines and enabled classification of dietary patterns in free-living populations (15). This strategy has been used to accurately assess dietary patterns in athletes (16) and obese (17) cohorts. However, it remains unclear whether this strategy developed in healthy adults can be used to establish dietary models within individuals with altered metabolic states, given that metabolic and inflammatory conditions are known to influence urinary metabolite composition (18–20). Extending dietary metabolic phenotyping modelling to individuals at increased CVD risk (e.g., elevated blood pressure, dyslipidemia, and with overweight) is therefore a critical step towards improving objective dietary assessment and for developing precision nutrition approaches in populations most likely to benefit.

We hypothesized that 72 hours (72 h) of full adherence to a NICE-compliant diet versus a Western-style dietary pattern would elicit distinct modulation of metabolic phenotypes in participants at elevated risk of CVD, and that using the same framework previously applied in heathy adults (15), would allow us to build a specific urinary metabolic phenotyping model to optimize the dietary assessment in this higher-risk group.

## Methods

This study was an inpatient randomized cross-over clinical trial conducted at UK National Institute for Health Research (NIHR) Imperial Clinical Research Facility. The study was approved by the London-Dulwich Research Ethics Committee Reference Number (REC Ref:18/LO/2042) and registered on the ISRCTN registry (Registration ID: ISRCTN44705179. https://www.isrctn.com/ISRCTN44705179). All participants provided written informed consent.

### Study participants

Participants were recruited from a database of volunteers at the NIHR Imperial Clinical Research Facility and through poster advertisement. Inclusion criteria were men and women of all ethnicities, aged between 30 and 65 years, who were at risk of CVD and met at least three of the following criteria (i) body mass index (BMI) of ≥ 25 and < 35 kg/m^2^; (ii) systolic blood pressure (SBP) ≥ 140 mmHg or diastolic blood pressure (DBP) ≥ 90 mmHg or current use of antihypertensive medication; (iii) Low density lipoprotein cholesterol (LDL-C) ≥ 4.14 mmol/l (≥ 160 mg/dl) and high-density lipoprotein cholesterol (HDL-C) ≤ 1.03 mmol/l (≤ 40 mg/dl) (men) or ≤ 1.29 mmol/l (≤ 50 mg/dl) (women); (iv) family history of premature coronary heart disease; (v) waist circumference > 102 cm in men or > 88 cm in women. Exclusion criteria included (i) weight change of ≥ 3 kg in the last three months; (ii) taken prescription medicines having an impact on metabolism, appetite regulation and hormonal regulation; (iii) taken any dietary supplements in the last six months; (iv) any chronic illness or being diagnosed with HIV; (v) any gastrointestinal disorder e.g. Crohn’s disease, coeliac disease, or irritable bowel syndrome; (vi) a history of drug or alcohol abuse in the last two years; (vii) pregnancy; (viii) involved in any other research studies during the past 12 weeks.

### Study design

Each participant made two visits to the clinical research facility, with each visit consisting of a 4-day inpatient stay. An outline of the study design is presented in **Figure 1**. During each study visit, participants arrived at the clinical unit at 17:00 on Day 0 and consumed a reference dinner (**Table S1**). On Day 1, participants were assigned to either a NICE-compliant or Western-style diet and initiated the 72 h dietary intervention (09:00 on Day 1 to 09:00 on Day 4); diet details are described below, with menus provided in **Table S2**. There was a minimum 14-day gap between visits to avoid possible carryover effects of the previous diet. Participants did not leave the clinical unit for the duration of the trial and consumed all food provided. Physical activity was low during the study visit.

**Figure 1.**
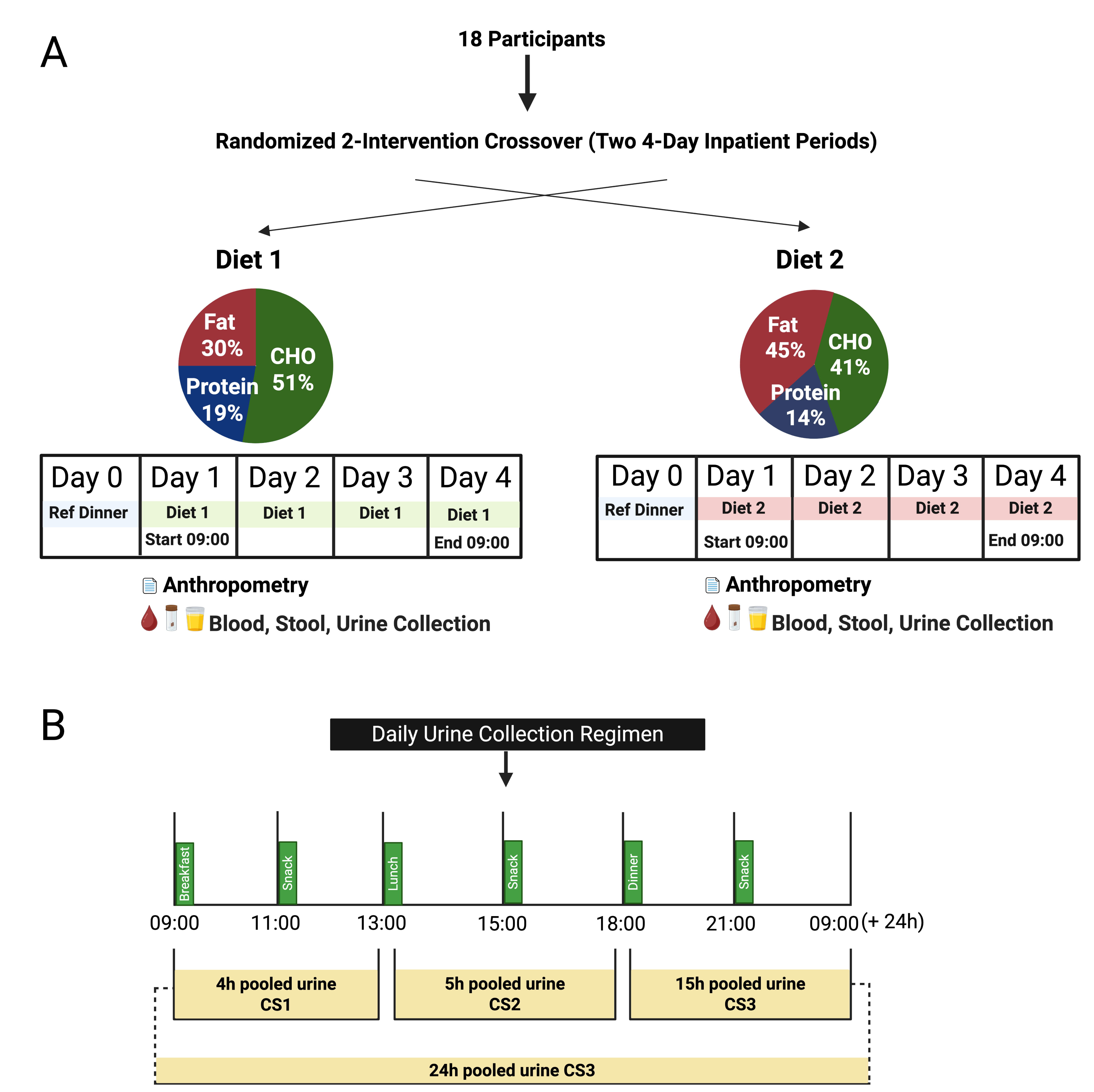
Study design. This was an inpatient randomized crossover dietary study in participants at risk of cardiovascular diseases (n=18). Each participant completed two 4-day inpatient visits separated by at least 14 days washout period. On Day 0 of each visit, participant arrived at the clinical research unit at 17:00 and were given a reference dinner before starting the dietary intervention the following morning. Participants received the dietary intervention from 09:00 on Day 1 to 09:00 on Day 4, which constituted a 72 hours (72 h) inpatient feeding period. During each visit, participant consumed either Diet 1 or Diet 2 from Day 1 to Day 4. Diet 1 followed guidelines established by the National Institute for Health and Care Excellence (NICE). Diet 2 represented a typical Western dietary pattern. The pie charts demonstrate the macronutrient proportions of protein, carbohydrates (CHO) and fat for each diet. After completing the first diet and a washout period of at least 14 days, participants crossed over to the alternate diet. On Day 1 and Day 4, morning (overnight-fasted) anthropometric measurements were recorded and fasting blood samples were collected. Stool samples were collected daily where possible. Urine was collected over three cumulative intervals (09:00–13:00 h, 13:00–18:00 h, and 18:00–09:00 h) during each 72 h inpatient stay, and pooled to obtain 24 h samples.

### Dietary Interventions

Two dietary interventions were designed: a NICE-compliant diet adhered to the UK NICE guidelines for cardiovascular disease prevention (21), and a Western-style diet reflecting a Western dietary pattern with minimal NICE concordance. Both interventions were isocaloric with identical total energy intake but differing macronutrient composition.

The macronutrient framework of NICE-compliant diet comprised carbohydrate at 45-55% of total energy intake, protein at 15-20%, and total fat at 25-29%. Specific targets within the carbohydrate intake comprised fibre (> 30 g daily) and free sugar (< 5% of total energy). Within the fat component, saturated fat was targeted at < 7% of total energy. Food sources were designated to include fruits and vegetables (> 400 g daily); fish (> 40 g daily), unsalted nuts and seeds (20 g daily) and baked beans (30.4 g daily); red meat intake was limited to < 70 g daily. Monounsaturated fats from olive oil were used for food preparation. The macronutrient framework for Western-style diet was derived from the National Diet and Nutrition Survey (NDNS) rolling programme data collected between 2016 and 2019 for UK adults aged 19-64 years (22). The NDNS provides representative data on dietary patterns in the UK adult population. The lowest observed dietary intakes (2.5^th^ and 16^th^ percentiles) of protein, fibre, and fruits and vegetables, and the highest observed intakes (84^th^ and 97.5^th^ percentiles) of total fat, saturated fat, trans fat, free sugars, salt, and red meat were used to represent a Western-style diet. The Western-style diet comprised carbohydrate contributing 37-44%, protein 11-13%, and total fat 42-48% of total energy intake. Within the carbohydrate intake, fibre content was 8-12 g daily and free sugar intake 85-143 g daily (15.5-23% of total energy). Within the fat component, saturated fat contributed 16-20% of total energy. Fruit and vegetable intake was 26-70 g daily, with no fish, nuts, seeds, or legumes included. Red meat consumption was 107-183 g daily, with refined grains as the main carbohydrate sources and no monounsaturated fat from plant oils. Dietary cholesterol in Western-style diet was > 300 mg daily and salt intake 12.5-17.8 g daily. Individual macronutrient composition designed for each participant was calculated based on their resting energy expenditure multiplied by a physical activity factor of 1.2 and details are provided in **Tables S3 and S4**.

### Randomization and masking

Eligible participants were randomly assigned to the order of the dietary intervention using a web-based sealed envelope randomization service provided by Sealed Envelope (www.sealedenvelope.com).

### Anthropometric measurements

Body composition was measured at baseline (Day 1) and post-intervention (Day 4, after 72 h) following an overnight fast after emptying the bladder, with participants wearing lightweight clothing. Anthropometric measurements including body weight, fat (%, kg), body water (%, kg), muscle (%, kg), and basal metabolic rate (BMR) were obtained using a body composition analyzer (Tanita, BC-418). Waist circumference was measured using a soft tape at the middle point between the lowest rib and the top of the hip bone, at the level of the belly button. Blood pressure was measured after participants had been seated at rest for at least 5 minutes in a relaxed state, with feet flat on the floor and arm supported (SureSigns@ VM4 patient monitor Philips). Participants were instructed not to talk during this time. SBP and DBP were measured three times, and the average was recorded.

### Assessment of cardiometabolic and glycoprotein signals

Fasting serum sample collected at baseline and after 72 h of adherence to dietary interventions were analyzed for glucose, triglyceride, cholesterol, LDL-C, HDL-C, LDL phospholipids (LDL-Phos), HDL-PL, apolipoprotein A1 (ApoA1), Apo-B100 and the glycoprotein signal (Glycs), α1-acid glycoprotein A (GlycA), α1-acid glycoprotein B (GlycB) were measured using Bruker PhenoRisk PACS^TM^ methods (23–24). Six LDL subfractions with increasing density (LDL-1 to LDL-6) were quantified using Bruker IVDr Lipoprotein Subclass Analysis B.I.LISA™ method (23).

### Global urine metabolic phenotyping using ^1^H-NMR spectroscopy

Urine was collected daily across three time periods: 09:00-13:00 h (morning collection; cumulative sample 1), 13:00-18:00 h (afternoon, cumulative sample 2), and 18:00-09:00 h (evening, overnight; cumulative sample 3). 24 h urine samples were obtained by pooling these three-time specific collections. The urine samples were prepared with a phosphate buffer following the previously described method (25). Urine samples were analyzed at 300 K on a 600 MHz spectrometer (Bruker BioSpin, Karlsruhe, Germany) utilizing a standard one-dimensional pulse sequence with water resonance saturation comprising: RD – gz,1 – 90° – t – 90° – tm – gz,2 – 90° – ACQ (25). RD is the relaxation delay of 4 s, t is a short delay of 4 μs, 90° is a 90° radio-frequency pulse, tm represents the mixing time of 10 ms, gz,1 and gz,2 are magnetic field z-gradients both applied for 1 ms, and ACQ is the data acquisition period of 2.7 s. The receiver gain was set to 90.5 in all experiments. Water suppression was achieved by continuously irradiating the sample with a 25Hz radio-frequency pulse at the water resonance frequency during the RD and tm stages. Each urine spectrum was obtained through 4 dummy scans, followed by 32 scans with 64K time domain points and with a spectral window set to 20 ppm for urine. Before performing the Fourier transformation, the acquired data were multiplied by an exponential function applying a line broadening factor of 0.3 Hz. The ^1^H-NMR spectra were manually phased and digitized over the range δ −0.5 to 9.5 and imported into MATLAB (Release 2019b). Each spectrum was baseline corrected using in-house software. The spectra were then referenced to the internal chemical shift reference (trimethylsilyl-[2,2,3,3-^2^H_4_]-propionate, TSP) at δ 0.0. Spectral regions containing the signals from the internal standard (δ −0.5 to 0.5) and water (δ 4.5 to 5.5) were excluded prior to probabilistic quotient normalization (26). After pre-processing, each spectrum consisted of 16,000 features.

### Global serum metabolic profiling using ^1^H-NMR spectroscopy

Fasting serum samples were prepared with a phosphate buffer following the previously described method (25). Serum samples were analyzed at 300 K on a 600 MHz Bruker Avance III HD spectrometer equipped with a 5 mm BBI probe and fitted with the Bruker SampleJetTM robot cooling system set to 5 °C. The following standard one-dimensional pulse sequence was used: RD – gz1 – 90° – t1 – 90° – tm – gz2 – 90° – ACQ (25). Relaxation delay (RD) of 4 s, interpulse delay (t1) was set to an interval of 4 μs, 90° is a 90° radio-frequency pulse, tm represent the mixing time of 10ms, gz,1 and gz,2 are magnetic field z-gradients both applied for 1 ms, and data acquisition period was 2.7 s. Water suppression was achieved by continuously irradiating the sample with a 25 Hz radio-frequency pulse at the water resonance frequency during the RD and tm stages. Each spectrum was acquired using 4 dummy scans followed by 32 scans. A spectral width of 12,000 Hz was used for all the samples. Prior to Fourier transformation, the free induction decays (FIDs) were multiplied by an exponential function corresponding to a line broadening of 0.3 Hz. Carr-Purcell-Meiboom-Gill (CPMG) one dimensional pulse sequence was used to attenuate broad, interfering peaks from lipids and proteins present in serum. The CPMG pulse sequence had the form RD – 90° – (t – 180° – t) n – ACQ. The acquisition parameters were set using the same settings as the standard 1D pulse sequence, with the spin-echo delay (t) set at 0.3 ms and 128 loops (n) performed. During the RD, continuous wave irradiation was applied at the water resonance frequency. The ^1^H-NMR spectra underwent automated adjustments to correct for phase and baseline distortions. They were also referenced to the TSP singlet at δ 0.0 and processed using TopSpin 3.1 software. Next, the spectra were digitized into 20K data points with a resolution of 0.0005 ppm using a custom MATLAB R2021b script. Sections of the spectra corresponding to the internal standard (δ −0.5 to 0.5) and water (δ 4.6-5) peaks were removed. To establish consistency, all spectra were normalized using median fold change normalization, using the median spectrum as the reference (26).

### Urinary and serum metabolite identification

A combined approach of data-driven and 2D NMR experiments were implemented to assist the structural identification of significant discriminatory metabolites (27). Briefly, the SubseT Optimization by Reference Matching (STORM) was used for data driven metabolite identification of the significant (q≤ 0.01) ^1^H-NMR variables (28). Chemical shifts were matched to metabolites using in-house library built with commercial chemical standards. The analytical strategies, specifically, a catalogue of ^1^H-NMR sequence with water pre-saturation and 2D NMR experiments such as J-Resolved spectroscopy, ^1^H-^1^H TOtal Correlation SpectroscopY (TOCSY), ^1^H-^1^H COrrelation SpectroscopY (COSY), ^1^H-^13^C Hetero-nuclear Single Quantum Coherence (HSQC) and ^1^H-^13^C Hetero-nuclear Multiple-Bond Correlation (HMBC) spectroscopy were performed. Chemical shifts were matched to metabolites using in-house library built with commercial chemical standards. Final metabolite identification was performed by spiking chemical standards when commercially available.

### Short chain fatty acid analyses

Short chain fatty acids (SCFAs) and the related metabolites, including lactate, acetate, 2-hydroxybutyrate, propionate, isobutyrate, butyrate, 2-methylbutyrate, isovalerate, valerate, and hexanoate, were quantified using liquid chromatography with electrospray ionization tandem mass spectrometry (LC-MS/MS) following published methods (29).

### Metagenomics data processing and taxonomic profiling

All metagenomic processing was conducted using the Teagasc high-performance computing cluster. Initially, all paired-end reads were subjected to quality control (QC) procedures through a validated pipeline (https://github.com/SegataLab/preprocessing), consisting of three subsequent steps : i) discarding of low-quality (quality<20), short (L<75bp) and with too many ambiguous nucleotides (N>2) reads using Trim Galore v0.6.6 (https://www.bioinformatics.babraham.ac.uk/projects/trim_galore/); ii) removal of human (HG19 human genome release) and bacteriophage phiX174 DNA (Illumina spike-in) contamination by mapping reads against their reference genomes through BowTie2 v2.2.9 (with parameter – sensitive-local) iii) Reads passing the filters were then sorted and split reads into forward, reverse and unpaired files. Cleaned-up reads were used for short read based taxonomic profiling. Compositional read mapping-based profiles were generated using MetaPhlAn 4 v4.1.1 using the mpa_vOct22_CHOCOPhlAnSGB_202403 database. The scripts for the bioinformatics tools described above were generated utilizing the resources available at the GitHub repository: https://github.com/SegataLab/MASTER-WP5-pipelines. These scripts were derived and adapted from the repository’s collection. Community-level microbiome analysis was carried out with hillR (v0.5.2) (30) to compute alpha and beta diversity values.

### Statistical analysis

#### Power and sample size

The sample size is based on the mathematical model from (15). Effect size estimation was based on the urinary excretion of hippurate as a result of increasing fruit and vegetable intake. In a highly control environment, an increase from 100 g to 300 g of fruits and vegetable intake resulted in a rise of 3.48 mmol/24 h in urine hippurate, with an SD 4.52 mmol/24 h. The resulting effect size is 0.772, with an alpha of 0.05 and power of 0.90, it requires at least 16 volunteers (based on a one tailed difference between two dependent (paired) means). Allowing for a drop-out of 20%, the target recruitment was set at 20 volunteers.

### Data analysis

#### Univariate analysis

In the analysis of paired samples, a paired t-test as performed if data were normally distributed, or a Wilcoxon signed rank test (31) was used for non-parametric data. For unpaired comparisons between two independent groups, an independent samples t-test was used for normally distributed data, or a Mann-Whitney U test was applied for non-parametric data. For categorical variables, chi-square test was employed. The p-values are adjusted for the false discovery rate q-value using the Storey-Tibshirani approach (32). A p-value and q-value of 0.05 or less as significance cut-off points ensures statistical rigor and reduces the chance of false positives. These analyses were performed using Python (v 3.12) and MATLAB (2019b, MathWorks, Natick, U.S.A).

#### Multivariate analysis

Changes in global serum profile were modelled using Repeated Measures Partial Least Squares Discriminant Analysis (RM-PLS-DA) in a Monte Carlo Cross-Validation (MCCV) framework to potentially identify distinctive signatures between diets, as described previously in Garcia-Perez et al. (15). From this model, the p-values were derived for specific metabolites. These p-values were then adjusted for multiple testing by calculating the false discovery rate q-value using the Storey-Tibshirani approach. All analyses were performed using MATLAB (2019b, MathWorks, Natick, U.S.A).

## Results

### Recruitment and participant characteristics

Between 12 April 2021 and 6 December 2021, a total of 457 individuals were assessed for eligibility; 306 did not meet the inclusion criteria and 131 declined participations after receiving further study information. Twenty participants were randomized. Two participants withdrew during the study, and 18 participants completed the clinical trial (**Figure 2**, CONSORT diagram).

**Figure 2.**
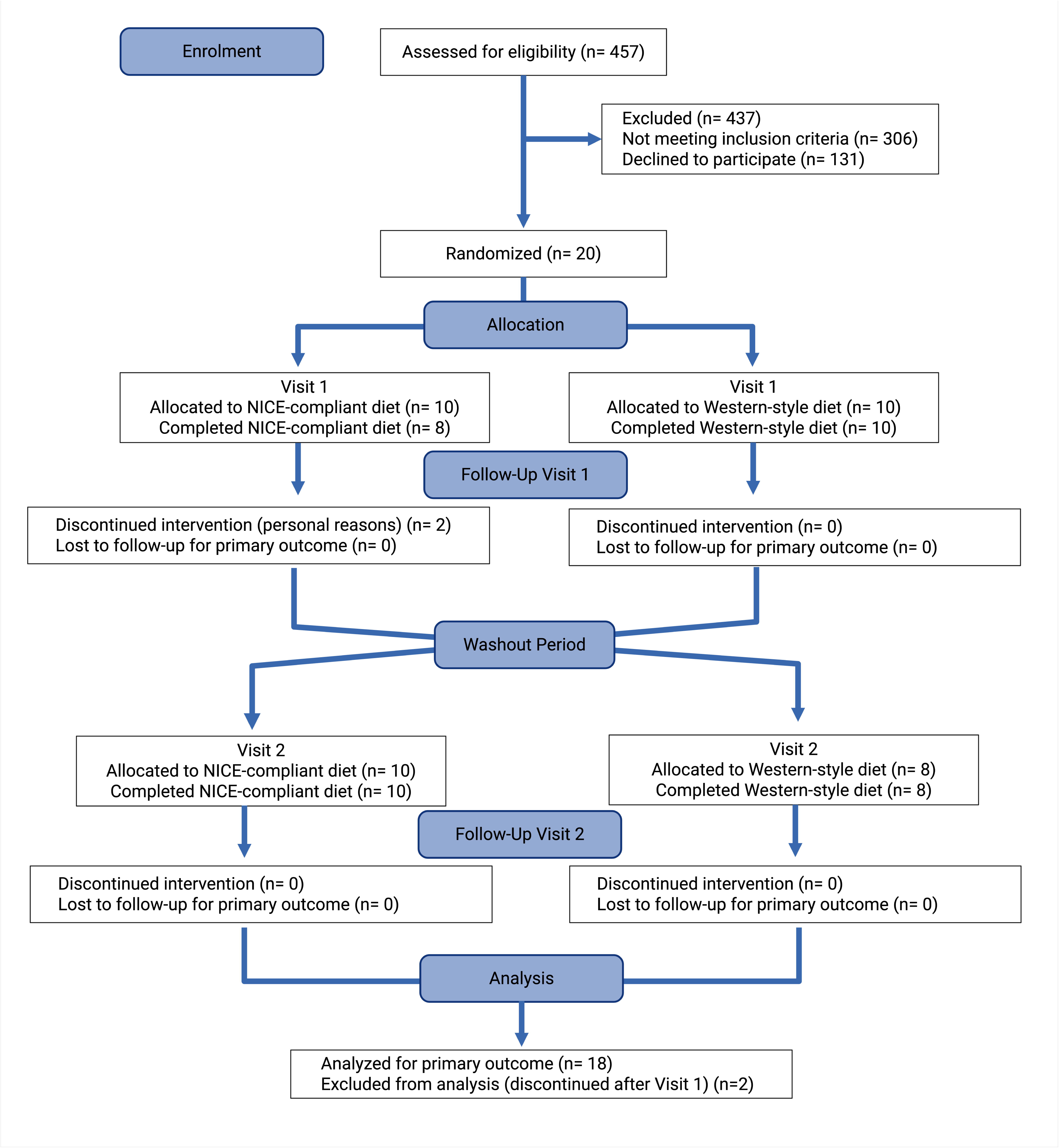
CONSORT 2025 flow diagram. Abbreviation: NICE, National Institute for Health and Care Excellence.

Demographic and baseline characteristics of the 18 participants who completed the crossover dietary intervention are presented in **Table 1**. The gender distribution of the participants was equal, and ethnicity was predominantly White (65%). Participants exhibited a CVD risk profile, with a median BMI of 30.6 kg/m^2^ in the overweight or obese range (30.6 kg/m^2^, IQR 28-31.6). Waist circumference (104.5 cm, 100.5-108.8) fell within the range of central obesity. Fasting glucose concentrations were within the normal range (5 mmol/L, 4.9-5.2). HDL-C concentrations was relatively low (45 mg/dL, 40.5-52.9). Baseline serum C-reactive protein (CRP) concentrations (1.1 mg/L, 0.7-1.9) were not indicative of systemic inflammation.

**Table 1.** Baseline characteristics of the study participants (n=18)

|  |  |
| --- | --- |
| Gender |  |
| Female | 9 (50%) |
| Male | 9 (50%) |
| Ethnicity |  |
| White | 13 (65%) |
| Black | 2 (10%) |
| Asian | 3 (15%) |
| Mixed ethnicity | 2 (10%) |
| Age | 54 (44-58) |
| Body composition |  |
| BMI (kg/m<sup>2</sup>) | 30.6 (28.0-31.6) |
| Fat (%) | 28.2 (24.8-40.2) |
| Body water (%) | 49.7 (42.7-53.1) |
| Muscle (%) | 68.2 (56.8-71.6) |
| BMR (kcal) | 1771 (1496-2081) |
| Waist circumference (cm) | 104.5 (100.5-108.8) |
| SBP (mmHg) | 126.2 (112.7-131.1) |
| DBP (mmHg) | 74.3 (69.2-77.7) |
| Glycaemic measures |  |
| HbA1c (mmol/mol) | 36.0 (33.8-37.0) |
| Glucose (mmol/l) | 5.0 (4.9-5.2) |
| Lipids and lipoproteins |  |
| Triglyceride (mg/dL) | 106.5 (89.5-122.9) |
| Cholesterol (mg/dL) | 173.5 (141.5-217.1) |
| LDL-C (mg/dL) | 87.3 (76.3-113.1) |
| HDL-C (mg/dL) | 45.0 (40.5-52.9) |
| LDL-PL (mg/dL) | 51.8 (45.3-64.4) |
| HDL-PL (mg/dL) | 64.0 (55.4-73.3) |
| Apo-A1 (mg/dL) | 123.8 (114.0-140.8) |
| Apo-B100 (mg/dL) | 76.5 (62.3-87.3) |
| Apo-B100/Apo-A1 | 0.5 (0.5-0.7) |
| Inflammation and glycoprotein signals |  |
| CRP (mg/L) | 1.1 (0.7-1.9) |
| GlycA (p.d.u) | 0.8 (0.7-0.8) |
| GlycB (p.d.u) | 0.3 (0.3-0.3) |
| Glyc (p.d.u) | 1.1 (1.0-1.1) |
| Data presented as Median (IQR) and n (%) for categorical variables. BMI, body mass index; BMR, basal metabolic rate; SBP, systolic blood pressure; DBP, diastolic blood pressure; HbA1c, haemoglobin A1c; LDL-C, low density lipoprotein cholesterol; HDL-C, high density lipoprotein cholesterol; LDL-PL, low density lipoprotein phospholipids; HDL-PL, high density lipoprotein phospholipids; Apo-A1, apolipoprotein-A1; Apo-B100, apolipoprotein-B100; CRP, C-reactive protein; Glycs, glycoprotein signal; GlycA, α1-acid glycoprotein A; GlycB, α1-acid glycoprotein B; p.d.u., procedure-defined units (NMR spectral units). | |

### Short term dietary adherence modulates CVD risk markers

Participants demonstrated a clear differential metabolic response to the two contrasting diets. SBP decreased after NICE-compliant but increased after the Western-style diet, resulting in a significant between diet difference (q = 7.2 x 10^-3^). No significant differences were observed between diets for body weight, BMI, body fat, body water, muscle mass, basal metabolic rate, waist circumference, or diastolic blood pressure (**Table S5**). Serum LDL-C, LDL phospholipids (LDL-PL), and the LDL-C:HDL-C ratio were significantly reduced after NICE-compliant diet, whereas these parameters increased following Western-style diet (q = 2.60 x 10^-2^, q = 2.60 x 10^-2^, and q = 1.30 x 10^-2^, respectively). Total cholesterol (q = 2.60 x 10^-2^) and Apo-B100 concentrations (q = 2.60 x 10^-2^) showed a significantly greater increase after Western-style diet (**Figure 3A, Table S6**). Density based stratification of LDL particles revealed that NICE-aligned diet selectively decreased both the particle number and lipid content of the larger, more buoyant LDL subclasses (LDL1-3). Significant decreases were observed in LDL1-3 particle number (LDL 1-3 PN, q =1.31 x 10^-2^), cholesterol (LDL 1-3 CH, q =1.31 x 10^-2^), free cholesterol (LDL 1-3 FC, q = 2.55 x 10^-2^), and phospholipids (LDL 1-3 PL, q =1.31 x 10^-2^). In contrast, these measures increased following Western-style diet (**Figure 3B, Table S7**). The smaller, denser, and more atherogenic LDL subclasses (LDL 4-6) were unaffected by either diet. Inflammatory markers, including CRP, NMR-derived GlycA and GlycB, and their composite signal (Glyc) were also showed no differential response to the dietary interventions (**Table S6**).

**Figure 3.**
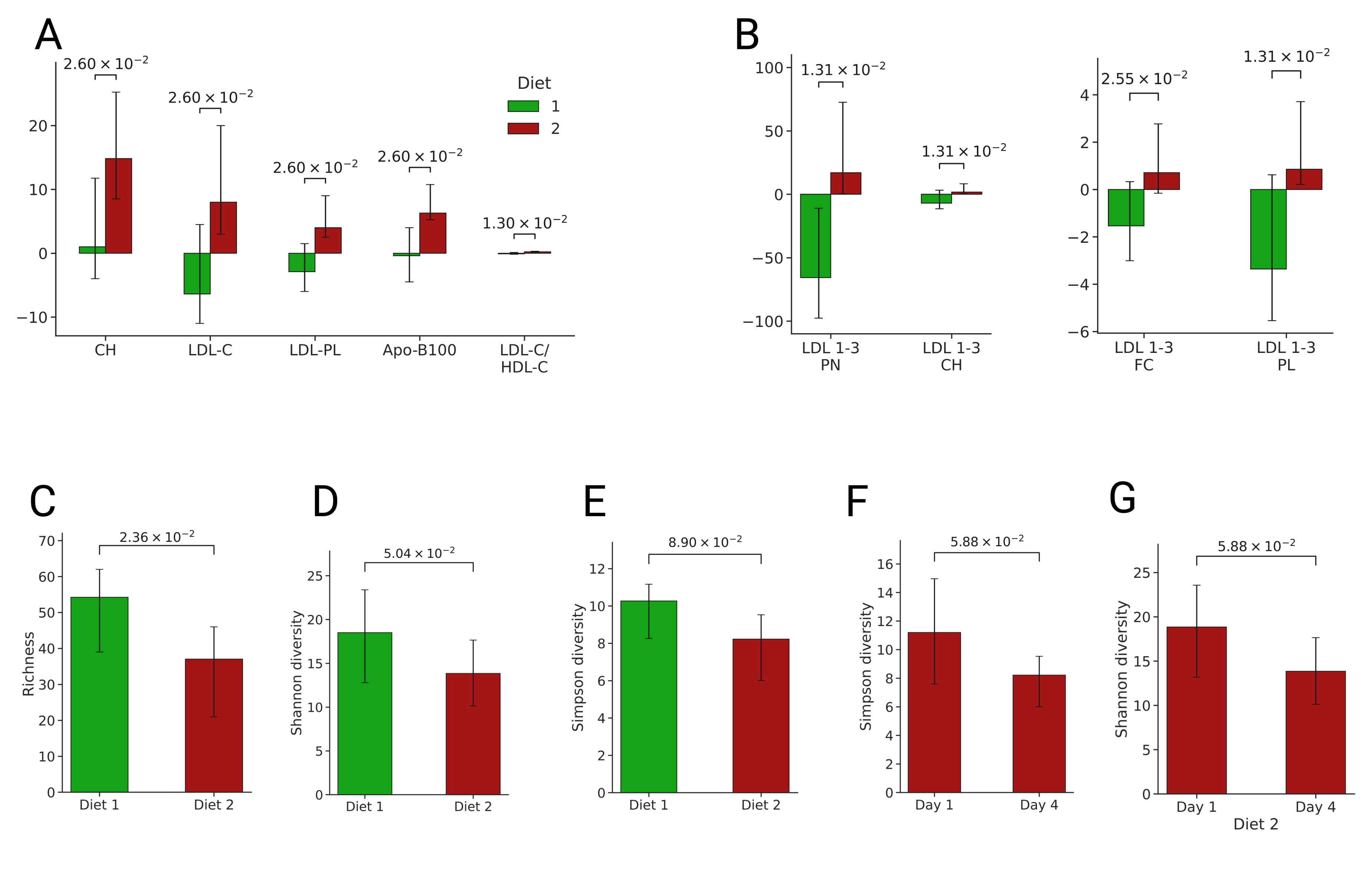
The impact of Diet 1 (NICE guidelines) compared with Diet 2 (Western) on serum lipids, lipoprotein subfractions and gut microbial alpha diversity measured from stool metagenomes (n=18). (A-B) Bar plots compare the impact (delta change: Day 4-Day 1) of Diet 1 and Diet 2 on lipids and lipoprotein and LDL subfractions. q-values are shown above the bars. (C) Microbial richness was significantly lower after Diet 2 compared to Diet 1 on Day 4 (after 72 h). (D-E) Shannon and Simpson diversity on Day 4 showed trends towards lower values after Diet 2 than Diet 1. (F-G) Within Diet 2, both Shannon and Simpson diversity showed trends toward a decrease from baseline to Day 4. Data are presented as median with interquartile range. p-values are shown above the corresponding bars. Diet 1 is shown in green; Diet 2 in red. Abbreviations: CH, Cholesterol; LDL-C, low density lipoproteins cholesterol; LDL-PL, low density lipoprotein phospholipids; Apo-B100, apolipoprotein-B100; PN, particle number; FC, free cholesterol; PL, phospholipids.

### Dietary adherence alters gut microbiome alpha diversity

Microbial richness was lower after Western-style diet compared with NICE-compliant diet (p = 2.36 x 10^-2^, **Figure 3C**). Moreover, Shannon and Simpson diversity showed trends towards lower diversity after Western-style diet (p = 5.04 x 10^-2^, p= 8.90 x 10^-2^ respectively; **Figure 3D-E**). Within diet analyses indicated no significant changes in microbial richness and diversity after NICE-compliant diet. In contrast, adherence to Western-style diet was associated with trends towards reduced diversity (p= 5.88 x 10^-2^, **Figure 3F-G**).

### Dietary adherence modulated serum and urinary metabolic phenotypes

Multivariate analysis using RM-MCCV-PLS-DA (R^2^Y = 0.96, Q^2^Y = 0.26) indicated rapid modulation of fasting serum low-molecular-weight metabolic phenotypes between dietary interventions (**Figure S1**). Discriminating metabolites present at significantly higher concentrations after the NICE-compliant diet, included valine (q = 5.31 x 10^-3^), dimethylglycine (q = 1.65 x 10^-5^), reflecting animal protein intake; trimethylamine-*N*-oxide (TMAO) (q = 1.25 x 10^-3^) associated with oily fish; and unsaturated lipids (q = 9.27 x 10^-3^) (**Tables S8**). Fasting serum acetate was also higher after NICE-compliant diet, as measured by both NMR (q = 9.70 x 10^-3^) and LC-MS (q = 3.25 x 10^-3^) (**Tables S8-9**). In contrast, alanine (q = 8.82 x 10^-3^), pyruvate (q = 9.07 x 10^-3^), and glycine (q = 9.18 x 10^-3^) exhibited higher levels following Western-style diet. SCFAs (propionate and butyrate), branched chain fatty acids (isobutyrate, isovalerate, and 2-methylbutyrate), the medium-chain fatty acid hexanoate, and other organic acids (lactate and 2-hydroxybutyrate) showed no significant between diet differences (**Tables S9**).

RM-MCCV-PLS-DA model (R^2^Y = 1, Q^2^Y = 0.96) of 24 h urine spectra revealed systematic differences between metabolic phenotype of NICE-compliant and Western-style, reflected in the Manhattan plot of the metabolic phenotypes (**Figure 4A**). From a total of 16,000 variables in the mean ^1^H-NMR spectrum, 14 identified metabolites (5034 features) were present in significantly higher concentrations in urine after 72 h of consumption of NICE-compliant compared with Western-style diet, while 13 metabolites were present in significantly higher after Western-style diet (**Table S10**). For example, urinary hippurate (a marker of fruit and vegetable intake; q = 4.15 x 10^-6^), tartrate (derived from grapes; q = 4.05 x 10^-10^), rhamnitol (associated with fruit intake; q = 1.41 x 10^-8^), proline betaine (citrus; q = 1.15 x 10^-8^), *N*-acetyl-*S*-methyl-cysteine sulfoxide (cruciferous vegetables; q = 3.31 x 10^-10^), TMAO (oily fish q = 2.1 x 10^-8^), and dimethylamine (fish; q = 3.74 x 10^-8^) were significantly higher after NICE-aligned compared with the Western-style diet. In contrast, urinary carnitine (red meat; q = 1.27 x 10^-7^), glucose (carbohydrates; q = 5.89 x 10^-6^), *N*-acetyl-*S*-(1Z)-propenylcysteine sulfoxide (onion; q = 3.7 x 10^-13^), the gut microbial metabolites phenylacetylglutamine (PAG) and 4-cresyl sulfate (4CS) (derived from bacterial fermentation of dietary aromatic amino acids; q = 5.16 x 10^-3^, q = 6.89 x 10^-6^) and signals corresponding to medium-chain fatty acids (C5-C10; q = 1.77 x 10^-23^) were higher following Western-style diet.

**Figure 4.**
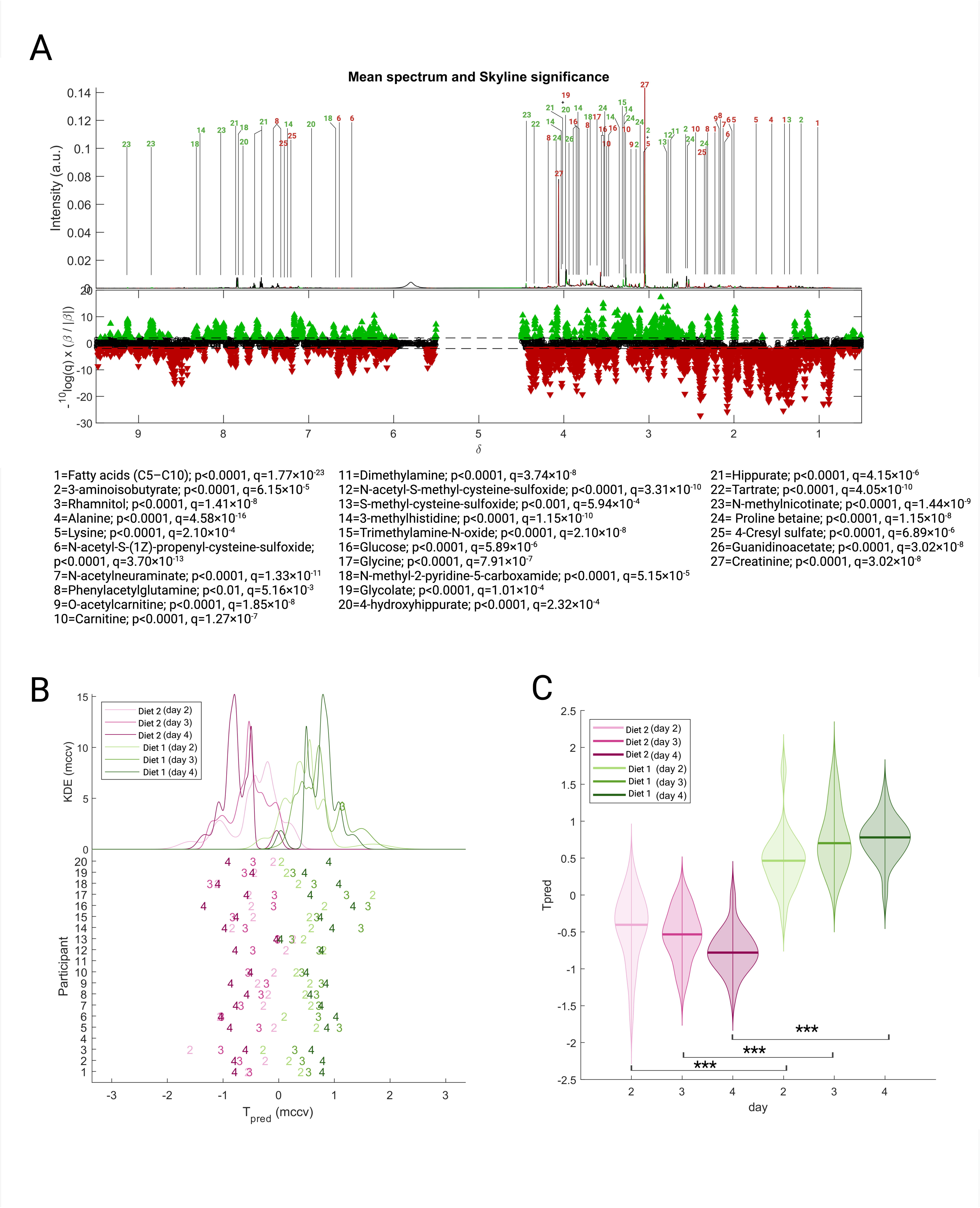
Urinary metabolic phenotyping discriminates between Diet 1 (NICE) and Diet 2 (Western). (A) Manhattan plot showing −log10(q) × sign of regression coefficient (β) of the MCCV-PLS-DA model for the 16,000 spectral variables. A p value was calculated for each variable on the basis of 25 bootstrap resamplings of the training data in each of 1,000 models to estimate the variance. Green peaks represent the 14 metabolites excreted in higher amounts after Diet 1 and red peaks represent the 12 metabolites excreted in higher amounts after Diet 2. The two horizontal lines indicate the cutoffs for the false discovery rate on the log_10_ scale. (B, C) Predicted scores from the MCCV-PLS-DA model, trained on using 24-hour urine metabolic phenotypes collected on Day 4 from the 18 participants following Diet 1 and Diet 2. These scores were used to predict 24 h samples from Days 2 and 3 for both diets in each participant. (B) The top part of the panel shows the Kernel Density Estimate (KDE) of the predicted scores (Tpred) for the four diets. The bottom part displays the Tpred from MCCV for each participant. (C) Violin plots of Tpred by day and diet. Pairwise comparisons between diets at each day were performed using Wilcoxon’s signed-rank test: Day 2, p = 7.63 × 10⁻⁶; Day 3, p = 7.63 × 10⁻⁶; Day 4, p = 7.63 × 10⁻⁶. MCCV-PLS-DA, partial least squares discriminant analysis model using Monte Carlo cross-validation.

The excretion time course of 30 metabolites quantified from the ^1^H-NMR spectra was quantified, based on measurements obtained after 24 h (Day 2), 48 h (Day 3), and 72 h (Day 4) of adherence to each dietary intervention. Concentrations of metabolites with well-established dietary associations, including hippurate (fruits and vegetable), proline betaine (citrus fruit), tartrate (grape), 1-methylnicotinate (plant-based protein), TMAO (fish, meat), dimethylamine (fish), 4-hydroxyhippuric acid (fruit), and valine (protein-rich foods) showed rapid increases on consumption of NICE-aligned diet, with significantly higher concentrations compared to Western style diet from 24 h onward (all q < 0.05, **Table S11**). In contrast, some metabolites required 72 h to manifest significant between-diet differences, including rhamnitol (plant-derived, apple), carnitine (red meat), and 1-methylnicotinamide (niacin rich foods) (all q < 0.05, **Table S11**). Urinary metabolites linked to gut microbial activity, such as hippurate (Hip) and the Hip: phenylacetylglutamine (PAG) ratio were significantly higher in NICE-aligned diet within 24 h and remained elevated throughout the intervention period (all q < 0.01, **Figure 5A,D**). Levels of 4-cresylsulfate, PAG, and the hippurate:4-cresyl sulfate (Hip:4CS) exhibited differences between diets after 48 hours (all q < 0.05, Figure 5 B,C,E). The PAG:4CS ratios showed a delayed response, with higher values in NICE-compliant diet observed at the end of the 72 h period (q = 2.14 x 10^-4^, **Figure 5F**).

**Figure 5.**
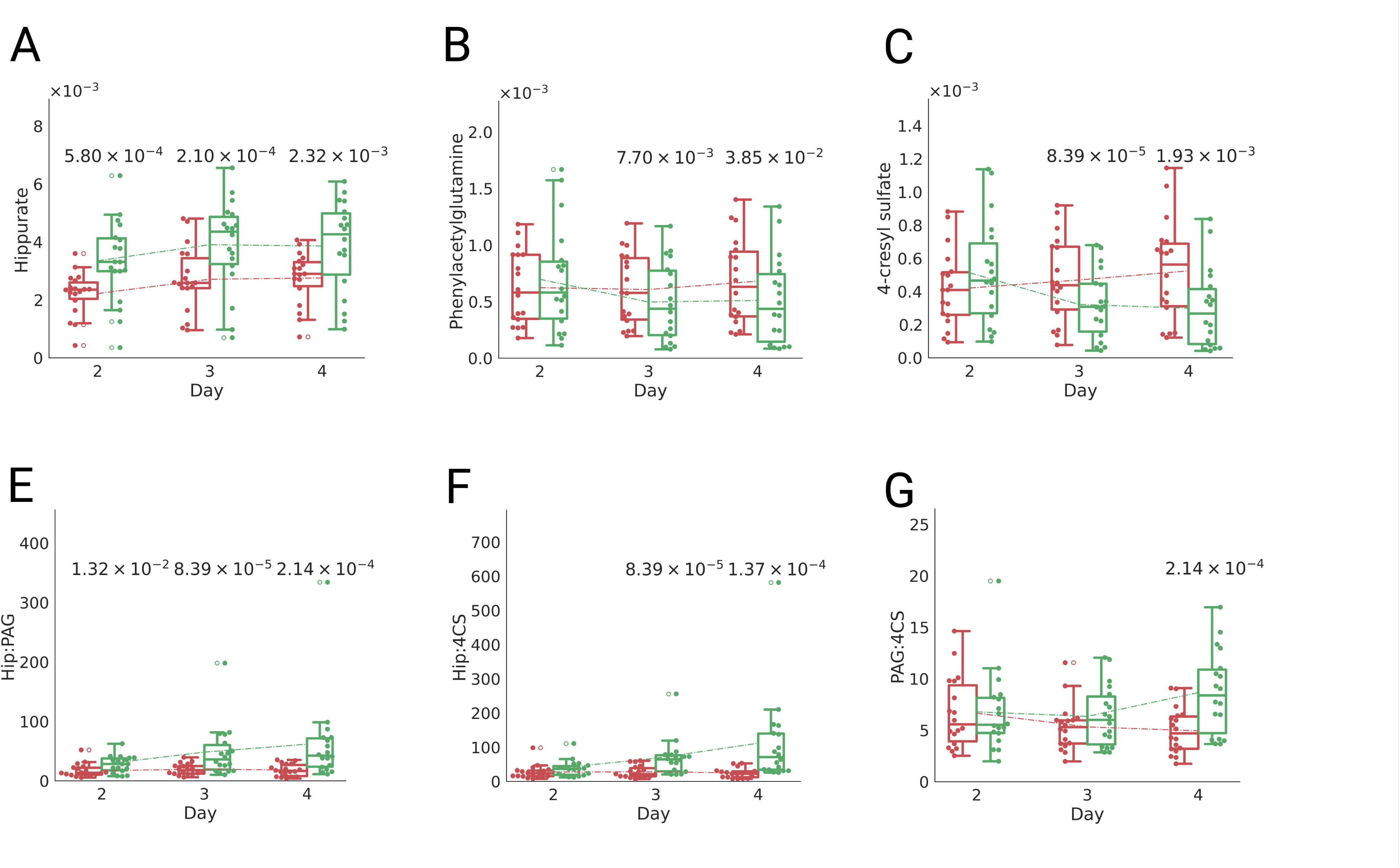
Temporal changes of gut microbial metabolites following Diet 1 (NICE) and Diet 2 (Western). (A) Hippurate (Hip), (B) Phenylacetylglutamine (PAG), (C) 4-cresyl sulfate (4CS), (D) Hip:PAG, (E) Hip:4CS, (F) PAG:4CS. Data in box plots are presented as median and interquartile range (IQR) with individual values representing measurements from each participant. Green and red indicate metabolite concentrations following Diet 1 and Diet 2, respectively. Statistical significance for C-H (q< 0.05) is indicated above corresponding time points with specific q-values.

### Assessing urinary metabolic phenotyping strategies for dietary assessment in CVD-risk participants

The methodology previously established by Garcia-Perez et al. (15) in healthy participants was applied to evaluate the ability of urinary metabolic phenotypes to assess dietary patterns in participants at risk of CVD, in accordance with NICE dietary guidelines. The RM-MCCV-PLS-DA model, previously developed using data from Day 4 (72 h) of the in-patient clinical trial, was validated using 24 h urinary metabolic profiles collected on Day 2 (24 h) and Day 3 (48 h) (**Figure 4B**). The predicted score (Tpred) derived from Day 4 model clearly separated two diets, with positive scores corresponding to NICE-compliant diet (clustered around Tpred≈1) and negative scores to Western-style diet (clustered around Tpred≈-1) (**Figure 4C**). Predicted scores on Days 2 and 3 progressively converged toward the Day 4 distributions, reflecting a consistent metabolic shift towards the diet specific profiles observed on Day 4 (**Figure 4C**). Pairwise comparisons between diets on Days 2, 3, and 4 revealed highly significant differences in predicted scores (all p < 0.001, **Figure 4C**).

## Discussion

Limitations in self-reported dietary assessment and incomplete adherence in free-living settings make it difficult to unravel the true effects of diet. An inpatient controlled feeding design offers a robust approach by ensuring all meals are provided and consumed under supervision, enabling precise assessment of dietary impact. Inpatient studies are constrained by cost and participant burden resulting in a relatively small sample size and short intervention periods. Tightly controlled inpatient feeding studies are resource intensive and therefore limited but have delivered strong experimental evidence on diet’s physiological impact (33–38). For example, Hall et al. (34) demonstrated that, under isocaloric energy restriction, restricting dietary fat resulted in greater net body fat loss than restricting carbohydrate. In a subsequent inpatient crossover trial, Hall et al. (35) reported that an ultra-processed diet led to significantly greater ad libitum energy intake and weight gain than an unprocessed diet matched for presented calories, macronutrients, and other nutrients (35).

Our results using an inpatient controlled feeding design in individuals at increased CVD risk, show that diet composition alone can modify CVD risk markers, in the absence of caloric restriction or changes in physical activity after 72 h full adherence. The NICE-compliant dietary pattern was associated with short-term reductions in SBP and atherogenic lipid measures including LDL-C relative to the Western-style diet. The two diets elicited changes in opposite directions, resulting in a marked between-diet contrast. Further investigation in LDL subfractions allowed for a more nuanced interpretation of dietary effects. Standard lipid panels do not capture LDL particle size, density or particles number, and smaller, denser LDL particles (sdLDL) are strongly associated with CVD risk (39, 40). Dietary effects on the LDL profile were driven mainly by the changes in larger LDL particles (LDL1 to LDL3), whereas sdLDL (LDL4 to LDL6) was not significantly modulated by diet. This may reflect the short intervention duration and the absence of a concurrent triglyceride response, as changes in sdLDL are often linked to alterations in triglyceride metabolism and insulin resistance (41).

In free living settings, dietary change can rapidly alter microbial community composition, but the reported effect on gut microbial richness and α-diversity has been inconsistent (42–46). In the current highly controlled study, we found that the Western-style diet was associated with lower gut microbial richness and a tendency towards reduced α-diversity compared with the NICE-compliant diet. This aligns with observational evidence linking Westernized or lower-quality dietary patterns characterized by high saturated fat and low fiber intakes with reduced gut microbiome diversity (47–49). In contrast, in a prior inpatient study, richness and α-diversity was not found to differ between Western and microbiome-enhancer diets (38), suggesting that these measures do not always shift over short dietary interventions in healthy adults (n=17) and therefore baseline metabolic status could be one contributor to the differing diversity responses.

Global metabolic phenotypes differed systematically between diets in both fasting serum and 24 h urine. The dietary intervention had a stronger impact on the urinary metabolic phenotype than on fasting serum, with a larger number of discriminatory metabolites capturing a broader range of food types and food groups, whereas the fasting serum profile showed a more restricted set of differences involving amino acids and lipid related metabolites. Urinary metabolite profiles are often more sensitive to dietary perturbations than circulating measures because systemic homeostatic regulation tightly constrains blood metabolite concentrations, whereas urine reflects the integrated excretion of transient dietary exposures, host-microbial co-metabolism, renal clearance dynamics, and short-term metabolic flux changes that are not buffered within the circulation. Discriminant urinary metabolites higher after the NICE-compliant diet included hippurate, tartrate (50), rhamnitol (51), and *N*-acetyl-*S*-methyl-cysteine sulfoxide (52), reflecting fruit and vegetable intake, as well as TMAO, dimethylamine, and 3-methylhistidines, reflecting fish and lean meat consumption (53). The Western-style diet was associated with increased urinary carnitine (54–55), glucose, and medium-chain fatty acids, consistent with greater red meat and refined carbohydrate consumption. In fasting serum, acetate was higher after the NICE-compliant diet, while other SCFAs were unchanged. A similar selective increase in fasting serum acetate has been reported following a diet compliant with WHO guidelines under the same 72 h inpatient setting (56), supporting the reproducibility of this response under controlled conditions. Higher fibre intake under guideline-compliant diets may contribute to acetate production via colonic fermentation in the fed state; however, fasting acetate also reflects endogenous metabolism alongside direct dietary sources (57), and therefore should not be interpreted as a specific marker of colonic fermentation.

We further characterized the progressive modulation of 30 urinary metabolites over the course of 72 h of adherence to each diet. Metabolites closely linked to specific foods and excreted largely unmetabolized, such as proline betaine and tartrate, increased by 24 h and remained elevated following the NICE-compliant diet. Metabolites produced through microbial and host metabolism, including 4-hydroxyhippuric acid and TMAO also increased over the first 24 h, consistent with previous interventions reporting rapid urinary excretion following polyphenol-rich fruit and fish intake respectively (58–59). In contrast, microbial host co-metabolites such as phenylacetylglutamine and 4-cresyl sulfate, derived from aromatic amino acid putrefaction of phenylalanine and tyrosine, increased more gradually and were highest at 48-72 h under the Western-style diet, reflecting a shift towards proteolytic fermentation in the distal colon likely driven by low dietary fiber intake (60). Higher Hip:PAG and Hip:4CS from 24 h onwards support a sustained predominance of saccharolytic fermentation in the proximal colon following the NICE-compliant diet (61).

The previously described metabolic phenotyping strategy (15) was applied to fasting serum and 24 h urine. The serum MCCV-PLS-DA model did not demonstrate sufficient robustness to reliably predict dietary intake (Q^2^Y = 0.26). In contrast, the 24 h urinary MCCV-PLS-DA model showed strong classification (Q^2^Y = 0.96), with the additional advantage of non-invasive sample. This performance underscores the robustness of the urinary metabolomic framework in detecting diet-induced changes. When urine samples collected on Days 2 and 3 were projected onto this model, they aligned with the expected diet-specific patterns observed after 72 h. The ability of this model to reproduce the expected direction of change at earlier time points demonstrates its capacity to capture the temporal trajectory of diet-induced metabolic shifts. Together, these findings support urinary metabolic phenotyping as a sensitive and non-invasive readout of diet-induced metabolic change and dietary exposure that complements traditional self-report methods for adherence assessment. This approach could be particularly valuable in individuals at elevated CVD risk, where accurate dietary assessment is central to both mechanistic and interventional prevention studies.

## Acknowledgements

IG-P, EH, and GF conceptualized the study. IG-P and GF designed the clinical trial, and LA and GF designed the isocaloric dietary interventions. GF and IG-P developed the methodology. JP and MK contributed software, data curation and analysis. YW, LA, and IG-P conducted the clinical trial. Formal analysis was performed by YW, JP, MK, LW, and IG-P; specifically, IG-P and JMP analyzed the metabolomics data, IG-P, YW, and MK analyzed the clinical trial data, OOS and LW conducted the metagenomic analyses, and IG-P performed metabolite identification. Visualization was performed by YW, JP, and MK. The original draft of the manuscript was written by YW, EH, GF, and IG-P. IG-P had primary responsibility for final content. All authors have read and approved the manuscript.

## Data Availability*

Data described in the manuscript, code book, and analytic code will be made available upon request pending [e.g., application and approval, payment, other]. Due to the small number of participants and potential for small numbers disclosure, data cannot be made publicly available. Any requests for (re-analysis of) data can be sent to the corresponding author. Contingent upon a data analysis plan and locally executable code, the corresponding author will re-analyse data locally and send summary-level results to requestors. All new code used to analyse the data has been shared on GitHub, with re-use of existing code referenced (including software and version number).

## Conflict of Interest

IG-P, JMP, EH and GF hold shares in Melico Sciences Ltd, IG-P, EH and GF are directors in the company. Melico has developed a quantitative method of assessing dietary intake using the same type of technology (NMR) as was used here to analyse urine samples. GF is lead for the Imperial Nestlé Collaboration and reports personal fees from Unilever, both outside the submitted work. All other authors declare no competing interests.

The author(s) declare that no generative AI or AI-assisted technologies were used in the writing of this manuscript.

## Abbreviations

^1^H-NMR: proton nuclear magnetic resonance spectroscopy;
4CS: 4-cresyl sulfate;
ACQ: data acquisition period;
Apo-A1: apolipoprotein A1;
Apo-B100: apolipoprotein B100;
BMI: body mass index;
BMR: basal metabolic rate;
CRP: C-reactive protein;
CVD: cardiovascular disease;
DBP: diastolic blood pressure;
GlycA: α1-acid glycoprotein A;
GlycB: α1-acid glycoprotein B;
Glycs: glycoprotein signal;
HDL-C: high-density lipoprotein cholesterol;
Hip: hippurate;
LDL-C: low-density lipoprotein cholesterol;
LDL-Phos: LDL phospholipids;
MCCV: Monte Carlo cross-validation;
NICE: National Institute for Health and Care Excellence;
NIHR: National Institute for Health Research;
PAG: phenylacetylglutamine;
RD: relaxation delay;
RM-PLS-DA: repeated-measures partial least squares discriminant analysis;
SBP: systolic blood pressure;
SCFAs: short chain fatty acids;
sdLDL: small dense low density lipoprotein;
TMAO: trimethylamine N-oxide;
Tpred: predicted score;
WHO: World Health Organization.

