## Supplementary Figures and Tables for "Metabolic Phenotyping Objectively Captures Dietary Intake and Short-term Cardiovascular Disease Risk Responses Under an Inpatient Randomized Crossover Clinical Trial"

### Supplementary Tables

### Supplementary Table 1. Dietary information of the reference meals.

| Meal description | Food Ingredients | Portion (per 100 g) | Energy (kcal) | Fat (g) | Saturated Fat (g) | Carbohydrate (g) | Sugars (g) | Fibre (g) | Protein (g) | Salt (g) |
|---|---|---|---|---|---|---|---|---|---|---|
| Penne pasta with chicken breast in a spiced tomato sauce | Cooked pasta (water, durum wheat semolina), tomato, chicken breast (15%), water, red pepper, tomato purée, onion, cornflour, olive oil, red chilli, salt, demerara sugar, garlic purée, balsamic vinegar (white wine vinegar, grape must concentrate), parsley, extra virgin olive oil, chilli powder, acidity regulators (citric acid, acetic acid), rapeseed oil. | 100 g | 119 | 1.7 | 0.4 | 16.7 | 3.3 | 1.9 | 8.3 | 0.33 |
| Conchiglie pasta with tuna in a tomato sauce topped with cheese sauce and mature cheddar cheese | Tomato, cooked pasta (water, durum wheat semolina), tuna (fish) (15%), water, whole milk, mature cheddar cheese (milk) (3%), cornflour, wheat flour (wheat flour, calcium carbonate, iron, niacin, thiamine), onion, tomato purée, butter (milk), garlic purée, salt, basil, sugar, black pepper, white pepper, nutmeg. | 100 g | 100 | 2.2 | 1.2 | 11.8 | 2.2 | 0.9 | 7.9 | 0.32 |

Participants chose either a chicken pasta or a tuna pasta reference dinner at visit 1 and received the same option at visit 2. They consumed the full pack (400 g) of their chosen meal at each study visit. Nutritional values are presented per 100 g as provided by the manufacturer (Tesco, UK).

**Supplementary Table 2. Representative dietary information and nutritional profiles of Diet 1 (NICE guidelines) and Diet 2 (Western), based on a standard energy intake of 2000 kcal/day.** Portion sizes were adjusted according to individual basal metabolic rates while maintaining identical food items across participants. This table illustrates a representative menu for Diet 1 and Diet 2.

| **Meal type**<br>(time) | **Breakfast** (09:00) | **Morning Snack** (11:00) | **Lunch**<br>(13:00) | **Afternoon Snack** (15:00) | **Dinner**<br>(18:00) | **Evening Snack**<br>(21:00) |
|---|---|---|---|---|---|---|
| Macronutrient composition | *Food (g or ml)* | *Food (g or ml)* | *Food (g or ml)* | *Food (g or ml)* | *Food (g or ml)* | *Food (g or ml)* |
| **Diet 1**<br>Proportion of protein: 17.4%<br>Proportion of carbohydrate: 50.6%<br>Total sugar: 120g<br>Proportion of fat: 27.4%<br>Saturated fatty acids: 5.2%<br>Monounsaturated fatty acids: 13%<br>Polyunsaturated fatty acids: 6.3%<br>Total trans fatty acids: 0.05%<br>Fibre: 46g<br>Sodium: 1177 mg<br>Fruit and vegetables: 1217g<br>Energy: 2004<br>Energy density: 0.62 | Banana (150 g)<br>Swiss Style Muesli (40 g)<br>Skimmed Milk (180 ml)<br>Tea (190g) | Orange (143 g)<br>Skimmed Milk (30 ml)<br>Coffee (190g) | Salmon and Dill Potato Bake (300g)<br>Mixed vegs (carrot, peas, cauliflower, cut green beans, sweetcorn) (200 g)<br>Egg Noodles (150 g)<br>Olive Oil (15 g)<br>Skimmed Milk (30 ml)<br>Tea (190g) | Grapes, red<br>(100 g)<br>Skimmed Milk (30 ml)<br>Coffee (190g) | Chicken Breast in Gravy<br>(165 g)<br>Jacket Potatoes<br>(114 g)<br>Baked Beans in Tomato sauce (57 g)<br>Mixed vegs (carrot, peas, cauliflower, cut green beans, sweetcorn)<br>(200 g)<br>Skimmed Milk (180 ml)<br>Tea (190g) | Apple (134 g)<br>Mixed nuts (25 g) |
| **Diet 2**<br>Proportion of protein: 13.5%<br>Proportion of carbohydrate: 42.4%<br>Total sugar: 84g<br>Proportion of fat: 43%<br>Saturated fatty acids: 21%<br>Monounsaturated fatty acids: 12%<br>Polyunsaturated fatty acids: 6%<br>Total trans fatty acids: 1.1%<br>Fibre: 10.5<br>Sodium: 3233<br>Fruit and vegetables: 30g<br>Energy: 2010<br>Energy density: 1 | Salted Butter (20 g)<br>White medium bread (40 g)<br>Whole Milk (180 ml)<br>Breakfast cereals, Cornflakes (40 g)<br>Tea (190g) | Chocolate Mousse (85 g)<br>Whole Milk (30 ml)<br>Coffee (190g) | Pork Sausages in Onion Gravy (250 g)<br>Mashed Potato (114 g)<br>Whole Milk (30 ml)<br>Tea (190g) | Chocolate<br>Bounty (28.5 g)<br>Whole Milk (30 ml)<br>Coffee (190g) | Quarter Pound Beef burger with Chargrilled onion (120 g)<br>Chips (102 g)<br>Burger Buns<br>(68 g) | Chocolate milk<br>(100 ml) |

### Supplementary Table 3. Individual energy requirements and corresponding macronutrient targets for Diet 1 (NICE guidelines).

| ID | Total energy (kcal/day) | Carbohydrate (45-55%) | | Protein (25-29%) | | Fat (25-29%) | | <7% Saturated fat (g) | Free sugar <5% (g) |
|---|---|---|---|---|---|---|---|---|---|
| | | Lower (g) | Upper (g) | Lower (g) | Upper (g) | Lower (g) | Upper (g) | | |
| 1 | 2737 | 307.91 | 376.34 | 102.64 | 136.85 | 76.03 | 88.19 | 21.29 | 36.49 |
| 2 | 2209 | 248.51 | 303.74 | 82.84 | 110.45 | 61.36 | 71.18 | 17.18 | 29.45 |
| 3 | 2500 | 281.25 | 343.75 | 93.75 | 125 | 69.44 | 80.56 | 19.44 | 33.33 |
| 4 | 1598 | 179.78 | 219.73 | 59.93 | 79.9 | 44.39 | 51.49 | 12.43 | 21.31 |
| 5 | 1744 | 196.2 | 239.8 | 65.4 | 87.2 | 48.44 | 56.2 | 13.56 | 23.25 |
| 6 | 2233 | 251.21 | 307.04 | 83.74 | 111.65 | 62.03 | 71.95 | 17.37 | 29.77 |
| 7 | 1750 | 196.88 | 240.63 | 65.63 | 87.5 | 48.61 | 56.39 | 13.61 | 23.33 |
| 8 | 1853 | 208.46 | 254.79 | 69.49 | 92.65 | 51.47 | 59.71 | 14.41 | 24.71 |
| 9 | 1927 | 216.79 | 264.96 | 72.26 | 96.35 | 53.53 | 62.09 | 14.99 | 25.69 |
| 10 | 1433 | 161.21 | 197.04 | 53.74 | 71.65 | 39.81 | 46.17 | 11.15 | 19.11 |
| 11 | 1920 | 216 | 264 | 72 | 96 | 53.33 | 61.87 | 14.93 | 25.6 |
| 12 | 1915 | 215.44 | 263.31 | 71.81 | 95.75 | 53.19 | 61.71 | 14.89 | 25.53 |
| 13 | 1776 | 199.8 | 244.2 | 66.6 | 88.8 | 49.33 | 57.23 | 13.81 | 23.68 |
| 14 | 2615 | 294.19 | 359.56 | 98.06 | 130.75 | 72.64 | 84.26 | 20.34 | 34.87 |
| 15 | 2462 | 276.98 | 338.53 | 92.33 | 123.1 | 68.39 | 79.33 | 19.15 | 32.83 |
| 16 | 2701 | 303.86 | 371.39 | 101.29 | 135.05 | 75.03 | 87.03 | 21.01 | 36.01 |
| 17 | 2540 | 285.75 | 349.25 | 95.25 | 127 | 70.56 | 81.84 | 19.76 | 33.87 |
| 18 | 2040 | 229.5 | 280.5 | 76.5 | 102 | 56.67 | 65.73 | 15.87 | 27.2 |
| 19 | 2494 | 280.58 | 342.93 | 93.53 | 124.7 | 69.28 | 80.36 | 19.4 | 33.25 |
| 20 | 1543 | 173.59 | 212.16 | 57.86 | 77.15 | 42.86 | 49.72 | 12 | 20.57 |

Diet 1 (NICE guidelines) were prepared for 20 enrolled participants according to individual energy requirements (see Figure 2, CONSORT diagram, for participant flow).

**Supplementary Table 4. Individual energy requirements and corresponding macronutrient targets for Diet 2 (Western).**

| | Total energy (kcal) | CHO (37-42%) | | Protein (11-13%) | | Fat (42-48%) | | Saturated fat (16%-20%) | | Free sugar (16%-23%) | |
|---|---|---|---|---|---|---|---|---|---|---|---|
| ID | | Lower (g) | Upper (g) | Lower (g) | Upper (g) | Lower (g) | Upper (g) | Lower (g) | Upper (g) | Lower (g) | Upper (g) |
| 1 | 2737 | 253.17 | 287.39 | 75.27 | 88.95 | 127.73 | 145.97 | 48.66 | 60.82 | 116.78 | 167.87 |
| 2 | 2209 | 204.33 | 231.95 | 60.75 | 71.79 | 103.09 | 117.81 | 39.27 | 49.09 | 94.25 | 135.49 |
| 3 | 2500 | 231.25 | 262.5 | 68.75 | 81.25 | 116.67 | 133.33 | 44.44 | 55.56 | 106.67 | 153.33 |
| 4 | 1598 | 147.82 | 167.79 | 43.95 | 51.94 | 74.57 | 85.23 | 28.41 | 35.51 | 68.18 | 98.01 |
| 5 | 1744 | 161.32 | 183.12 | 47.96 | 56.68 | 81.39 | 93.01 | 31 | 38.76 | 74.41 | 106.97 |
| 6 | 2233 | 206.55 | 234.47 | 61.41 | 72.57 | 104.21 | 119.09 | 39.7 | 49.62 | 95.27 | 136.96 |
| 7 | 1750 | 161.88 | 183.75 | 48.13 | 56.88 | 81.67 | 93.33 | 31.11 | 38.89 | 74.67 | 107.33 |
| 8 | 1853 | 171.4 | 194.57 | 50.96 | 60.22 | 86.47 | 98.83 | 32.94 | 41.18 | 79.06 | 113.65 |
| 9 | 1927 | 178.25 | 202.34 | 52.99 | 62.63 | 89.93 | 102.77 | 34.26 | 42.82 | 82.22 | 118.19 |
| 10 | 1433 | 132.55 | 150.47 | 39.41 | 46.57 | 66.87 | 76.43 | 25.48 | 31.84 | 61.14 | 87.89 |
| 11 | 1920 | 177.6 | 201.6 | 52.8 | 62.4 | 89.6 | 102.4 | 34.13 | 42.67 | 81.92 | 117.76 |
| 12 | 1915 | 177.14 | 201.08 | 52.66 | 62.24 | 89.37 | 102.13 | 34.04 | 42.56 | 81.71 | 117.45 |
| 13 | 1776 | 164.28 | 186.48 | 48.84 | 57.72 | 82.88 | 94.72 | 31.57 | 39.47 | 75.78 | 108.93 |
| 14 | 2615 | 241.89 | 274.58 | 71.91 | 84.99 | 122.03 | 139.47 | 46.49 | 58.11 | 111.57 | 160.39 |
| 15 | 2462 | 227.74 | 258.51 | 67.71 | 80.02 | 114.89 | 131.31 | 43.77 | 54.71 | 105.05 | 151 |
| 16 | 2701 | 249.84 | 283.61 | 74.28 | 87.78 | 126.05 | 144.05 | 48.02 | 60.02 | 115.24 | 165.66 |
| 17 | 2540 | 234.95 | 266.7 | 69.85 | 82.55 | 118.53 | 135.47 | 45.16 | 56.44 | 108.37 | 155.79 |
| 18 | 2040 | 188.7 | 214.2 | 56.1 | 66.3 | 95.2 | 108.8 | 36.27 | 45.33 | 87.04 | 125.12 |
| 19 | 2494 | 230.7 | 261.87 | 68.59 | 81.06 | 116.39 | 133.01 | 44.34 | 55.42 | 106.41 | 152.97 |
| 20 | 1543 | 142.73 | 162.02 | 42.43 | 50.15 | 72.01 | 82.29 | 27.43 | 34.29 | 65.83 | 94.64 |

Diet 2 (Western) were prepared for 20 enrolled participants according to individual energy requirements (see Figure 2, CONSORT diagram, for participant flow).

**Supplementary Table 5. The impact of Diet 1 (NICE guidelines) compared with Diet 2 (Western) on anthropometric measurements following 72-hour strict adherence (n=18).**

| | Diet 1 (Δ) | Diet 2 (Δ) | P-value | Q-value |
|---|---|---|---|---|
| Weight (kg) | -0.45 (-1.43--0.05) | -0.25 (-0.93-0.43) | $2.24 \times 10^{-1}$ | $8.79 \times 10^{-2}$ |
| BMI (kg/m2) | -0.15 (-0.50-0.00) | -0.10 (-0.53-0.20) | $9.36 \times 10^{-1}$ | $2.29 \times 10^{-1}$ |
| Fat (%) | 0.70 (-0.23-1.35) | -0.10 (-1.15-0.75) | $2.42 \times 10^{-1}$ | $8.79 \times 10^{-2}$ |
| Fat (kg) | 0.50 (-0.45-1.13) | -0.30 (-1.15-0.73) | $2.69 \times 10^{-1}$ | $8.79 \times 10^{-2}$ |
| Body Water (%) | -0.75 (-1.08-0.18) | 0.20 (-0.60-0.73) | $2.16 \times 10^{-1}$ | $8.79 \times 10^{-2}$ |
| Body Water (kg) | -0.70 (-1.43-0.05) | -0.25 (-0.73-0.80) | $2.02 \times 10^{-1}$ | $8.79 \times 10^{-2}$ |
| Muscle (%) | -0.70(-1.25-0.25) | 0.15 (-0.68-1.13) | $2.42 \times 10^{-1}$ | $8.79 \times 10^{-2}$ |
| Muscle (kg) | -0.75 (-1.43-0.05) | -0.35 (-0.95-1.15) | $1.85 \times 10^{-1}$ | $8.79 \times 10^{-2}$ |
| BMR (kcal) | -22.0 (-38.5--1.25) | -11.0 (-26.80-29.80) | $3.19 \times 10^{-1}$ | $9.39 \times 10^{-2}$ |
| Waist Circumference (cm) | 0.0 (-1.13-1.00) | 0.00 (-1.13-2.28) | $1.97 \times 10^{-1}$ | $8.79 \times 10^{-2}$ |
| SBP (mmHg) † | -2.85 (-13.05-2.78) | 1.85 (-3.08-8.45) | $2.44 \times 10^{-3}$** | $7.18 \times 10^{-3}$** |
| DBP (mmHg) | 0.35 (-3.40-4.30) | 1.85 (-3.40-8.30) | $5.87 \times 10^{-1}$ | $1.57 \times 10^{-1}$ |

Values presented as median (interquartile range).

Δ, Delta change (Day 4 - Day 1). Diet 1: NICE-compliant diet; Diet 2: Western-style diet.

† Significant variables fulfilling both p-value and FDR adjusted q-value thresholds of < 0.05.

*q<0.05, **q<0.01, ***q<0.001.

BMI, Body Mass Index; BMR, Basal Metabolic Rate; SBP, Systolic Blood Pressure; DBP, Diastolic Blood Pressure.

**Supplementary Table 6. The impact of Diet 1 (NICE guidelines) compared with Diet 2 (Western) on cardiometabolic risk factors following 72-hour strict adherence (n=18).** Lipid and lipoprotein measures showing significant between-diet differences after FDR correction (q<0.05) are presented in Figure 3.

| | Diet 1 (Δ) | Diet 2 (Δ) | P-value | Q-value |
|---|---|---|---|---|
| Glucose (mmol/l) | -0.1 (-0.2-0.2) | 0.0 (-0.3-0.2) | $6.40 \times 10^{-1}$ | $4.46 \times 10^{-1}$ |
| TG (mg/dL) | 13.0 (-8.8-26.0) | 6.5 (-8.8-36.2) | $4.88 \times 10^{-1}$ | $3.76 \times 10^{-1}$ |
| HDL-C (mg/dL) | -2.0 (-3.8-3.2) | 1.5 (-3.5-4.0) | $4.79 \times 10^{-1}$ | $3.76 \times 10^{-1}$ |
| HDL-PL(mg/dL) | 0.5 (-5.2-3.8) | 1.0 (-1.5-3.0) | $2.38 \times 10^{-1}$ | $2.21 \times 10^{-1}$ |
| Apo-A1 (mg/dL) | 0.0 (-4.5-6.8) | 3.5 (1.0-9.0) | $1.17 \times 10^{-1}$ | $1.38 \times 10^{-1}$ |
| ABA1 | 0.0 (-0.0-0.0) | 0.0 (0.0-0.1) | $9.20 \times 10^{-2}$ | $9.50 \times 10^{-2}$ |
| CH/HDL-C | 0.3 (-0.0-0.4) | 0.2 (0.1-0.4) | $7.40 \times 10^{-2}$ | $9.60 \times 10^{-2}$ |
| TG/HDL-C | 0.4 (-0.2-0.8) | 0.1 (-0.3-0.9) | $9.90 \times 10^{-1}$ | $5.36 \times 10^{-1}$ |
| GlycA (p.d.u) | 0.0 (0.0-0.1) | 0.0 (0.0-0.1) | $8.26 \times 10^{-1}$ | $5.36 \times 10^{-1}$ |
| GlycB (p.d.u) | 0.0 (0.0-0.0) | 0.0 (0.0-0.0) | $7.37 \times 10^{-1}$ | $5.07 \times 10^{-1}$ |
| Glyc (p.d.u) | 0.1 (0.0-0.1) | 0.0 (0.0-0.1) | $8.57 \times 10^{-1}$ | $5.36 \times 10^{-1}$ |
| CRP (mg/L) | -0.1 (-2-0.1) | -0.1 (-0.5-1.3) | $1.96 \times 10^{-1}$ | $2.00 \times 10^{-1}$ |

Values presented as median (interquartile range).

Diet 1: NICE-compliant diet; Diet 2: Western-style diet

† Significant variables fulfilling both p-value and FDR adjusted q-value thresholds of < 0.05.

*<0.05, **<0.01, ***<0.001 (for both p-values and q-values)

TG, triglycerides; HDL-C, high density lipoprotein cholesterol; HDL-PL, high density lipoprotein phospholipids; Apo-A1, apolipoprotein-A1; ABA1, Apo-B100/Apo-A1, Glycs, glycoprotein signal; GlycA, α1-acid glycoprotein A; GlycB, α1-acid glycoprotein B; CRP, C-reactive protein; p.d.u., procedure-defined units (NMR spectral units).

**Supplementary Table 7. The impact of Diet 1 (NICE guidelines) compared with Diet 2 (Western) on LDL subfraction particle number and lipid composition following strict adherence (n=18).** Lipoprotein measures showing significant between-diet differences after FDR correction (q<0.05) are presented in Figure 3.

| LDL subfraction | Diet 1 (Δ) | Diet 2 (Δ) | P-value | Q-value |
|---|---|---|---|---|
| LDL 1-3 TG | -0.03 (-1.54-2.02) | 0.08 (-0.63-0.94) | $5.98 \times 10^{-1}$ | $1.73 \times 10^{-1}$ |
| LDL 4-6 PN | 46.81 (-46.15-102.1) | 95.53 (32.9-138.7) | $2.28 \times 10^{-1}$ | $7.40 \times 10^{-2}$ |
| LDL 4-6 TG | 0.57 (-0.43-1.94) | 0.51 (-0.12-1.84) | $7.67 \times 10^{-1}$ | $1.99 \times 10^{-1}$ |
| LDL 4-6 CH | 0.74 (-4.98-5.53) | 7.86 (2.06-10.75) | $1.18 \times 10^{-1}$ | $4.37 \times 10^{-2}$ |
| LDL 4-6 FC | 0.41 (-1.67-1.28) | 1.64 (0.37-3.26) | $1.11 \times 10^{-1}$ | $4.37 \times 10^{-2}$ |
| LDL 4-6 PL | 0.24 (-2.08-3.11) | 3.70 (1.44-5.45) | $1.05 \times 10^{-1}$ | $4.37 \times 10^{-2}$ |

Values presented as median (interquartile range).
Diet 1: NICE-compliant diet; Diet 2: Diet 2: Western-style diet
† Significant variables fulfilling both p-value and FDR adjusted q-value thresholds of < 0.05.
*<0.05, **<0.01, ***<0.001 (for both p-values and q-values)
LDL subfractions are grouped as large LDL (LDL1-3, representing the sum of LDL1, LDL2, and LDL3) and small LDL (LDL4-6, representing the sum of LDL4, LDL5, and LDL6).
LDL, low density lipoprotein; PN, particle number; TG, triglycerides; CH, cholesterol; FC, free cholesterol; PL, phospholipids.

**Supplementary Table 8. List of serum metabolites with significant differences in concentrations between the Diet 1 (NICE guidelines) and Diet 2 (Western) measured by NMR after 72-hour strict adherence (n=18).** ↑ indicates higher excretion after Diet 1, a NICE-compliant diet; ↓ indicates higher excretion after Diet 2, the Western-style diet. s – singlet, d – doublet, t – triplet, q – quartet, dd – doublet of doublets, 2d – two doublets, m – (other) multiplet, b- broad. The table shows the significant variables fulfilling both p-value and FDR adjusted q-value thresholds of ≤ 0.05.

| Number | Metabolite | Chemical shift (multiplicity) | Dietary sources | Trend | P-value | Q-value |
|---|---|---|---|---|---|---|
| 1 | Alanine | 1.48 (d), 3.77 (q) | animal protein | ↓ | $1.67 \times 10^{-3}$ | $8.82 \times 10^{-3}$ |
| 2 | Pyruvic | 2.365 (s) | - | ↓ | $1.73 \times 10^{-3}$ | $9.07 \times 10^{-3}$ |
| 3 | Glycine | 3.57 (s) | Animal based protein | ↓ | $1.75 \times 10^{-3}$ | $9.18 \times 10^{-3}$ |
| 4 | Valine | 0.98 (d), 1.03 (d), 2.28 (m), 3.6 (d) | Whey protein dairy products | ↑ | $8.77 \times 10^{-4}$ | $5.31 \times 10^{-3}$ |
| 5 | Acetate | 1.91 (s) | Fibre | ↑ | $1.88 \times 10^{-3}$ | $9.70 \times 10^{-3}$ |
| 6 | Lipid CH=CH | 2.73 (b) | Unsaturated fat | ↑ | $3.34 \times 10^{-5}$ | $3.63 \times 10^{-4}$ |
| 7 | Dimethylglycine | 2.92 (s) | Egg and low-fat milk | ↑ | $8.35 \times 10^{-7}$ | $1.65 \times 10^{-5}$ |
| 8 | Trimethylamine-*N*-oxide | 3.25 (s) | Oily fish | ↑ | $1.46 \times 10^{-4}$ | $1.25 \times 10^{-3}$ |
| 9 | Unsaturated lipids ($CH=CHCH_2CH=CH$) | 5.33 (b) | Unsaturated fat | ↑ | $1.78 \times 10^{-3}$ | $9.27 \times 10^{-3}$ |
| 12 | 3-hydroxybutyric acid | 2.31(m), 2.38 (m) | - | ↑ | $1.19 \times 10^{-3}$ | $6.71 \times 10^{-3}$ |
| 14 | Ascorbate | 4.03 (dd), 4.52 (d) | Citrus fruits, leafy and root vegetables | ↑ | $1.83 \times 10^{-3}$ | $9.49 \times 10^{-3}$ |

**Supplementary Table 9. Comparison of serum short chain fatty acids (SCFAs) and the related metabolites after 72-hour strict adherence to Diet 1 (NICE guidelines) and Diet 2 (Western) (n=18).**

| Metabolite | Diet 1 | Diet 2 | P-value | Q-value |
|---|---|---|---|---|
| Lactate | 240.0 (233.1 - 264.07) | 235.6 (232.7 - 249.3) | 8.3 x 10-2 | 1.77 x 10-1 |
| Acetate † | 24.9 (15.87 - 34.83) | 10.5 (9.5 - 14.3) | 7.63 x 10-4 *** | 3.25 x 10-3 ** |
| Propionate | 1.9 (1.64 - 2.24) | 1.6 (1.1 - 1.8) | 2.11 x 10-1 | 2.46 x 10-1 |
| 2-hydroxybutyrate | 13.5 (11.42 - 16.58) | 9.7 (5.2 - 14.0) | 3.75 x 10-1 | 3.20 x 10-1 |
| Isobutyrate | 0.3 (0.23 - 0.4) | 0.3 (0.2 - 0.4) | 7.44 x 10-1 | 3.52 x 10-1 |
| Butyrate | 3.7 (3.19 - 4.26) | 3.1 (2.4 - 4.0) | 5.28 x 10-1 | 3.21 x 10-1 |
| 2-methylbutyrate | 0.1 (0.06 - 0.1) | 0.1 (0.0 - 0.1) | 5.28 x 10-1 | 3.21 x 10-1 |
| Isovalerate | 0.1 (0.12 - 0.3) | 0.1 (0.1 - 0.4) | 2.31 x 10-1 | 2.46 x 10-1 |
| Hexanoate | 0.3 (0.13 - 0.34) | 0.2 (0.1 - 0.4) | 7.06 x 10-1 | 3.52 x 10-1 |

Values presented as median (interquartile range).
Diet 1: NICE-compliant diet; Diet 2: Western-style diet
† Significant variables fulfilling both p-value and FDR adjusted q-value thresholds of < 0.05.
*<0.05, **<0.01, ***<0.001 (for both p-values and q-values).

**Supplementary Table 10. List of metabolites showing differences urinary excretion between Diet 1 (NICE compliant) and Diet 2 (Western).**

| | *Metabolite* | Chemical Shift (multiplicity)* | Association^ | *P-value/ Q-value* | *Dietary sources* |
|---|---|---|---|---|---|
| 1 | Fatty acids (C5-C10) | 0.88 (m), 1.31 (m), 2.19 (m) | ↓ | $2.08x10^{-26}$ $1.77x10^{-23}$ | Fats |
| 2 | 3-aminoisobutyrate | 1.19 (d), 2.6 (m), 3.02 (t), 3.09 (d) | ↑ | $2.17x10^{-5}$ $6.15x10^{-5}$ | Fruits |
| 3 | Rhamnitol | 1.28 (d) | ↑ | $1.91x10^{-9}$ $1.41x10^{-8}$ | Apple |
| 4 | Alanine | 1.48 (d) | ↓ | $7.06x10^{-18}$ $4.58x10^{-16}$ | Protein |
| 5 | Lysine | 1.73 (m), 1.91 (m), 3.02 (t) | ↓ | $8.54x10^{-5}$ $2.10x10^{-4}$ | |
| *6* | *N*-acetyl-*S*-(1Z)-propenyl-cysteine-sulfoxide | 1·96 (dd), 2·03 (s), 6·49 (dq), 6·65 (dq) | ↓ | $1.38x10^{-14}$ $3.70x10^{-13}$ | Onion |
| *7* | *N*-acetyl neuraminate | 2.06 (s) | ↓ | $7.81x10^{-13}$ $1.33x10^{-11}$ | |
| 8 | Phenylacetylglutamine | 2·11 (m), 2·27 (m), 3·67 (m), 4·19 (m), 7·36 (t), 7·43 (t) | ↓ | $3.08x10^{-3}$ $5.16x10^{-3}$ | Animal protein |
| *9* | *O*-acetylcarnitine | 2·15 (s), 3·19 (s) | ↓ | $2.57x10^{-9}$ $1.85x10^{-8}$ | (Red) meats |
| 10 | Carnitine | 2·44 (dd), 3·23 (s), 3·43 (m) | ↓ | $2.23x10^{-8}$ $1.27x10^{-7}$ | (Red) meats |
| 11 | Dimethylamine | 2.72 (s) | ↑ | $5.60x10^{-9}$ $3.74x10^{-8}$ | Fish |
| *12* | *N*-acetyl-*S*-methyl-cysteine-sulfoxide | 2.78 (s) | ↑ | $2.95x10^{-11}$ $3.31x10^{-10}$ | Cruciferous vegetables |
| *13* | *S*-methyl-cysteine-sulfoxide | 2.84 (s) | ↑ | $2.69x10^{-4}$ $5.94x10^{-4}$ | Cruciferous vegetables |
| 14 | 3-methylhistidine | 3·25 (2d), 3·30 (2d), 3·78 (s), 3·99 (dd), 7·23 (s), 8·27 (s) | ↑ | $9.15x10^{-12}$ $1.15x10^{-10}$ | (Lean, white) meats |
| 15 | Trimethylamine-*N*-oxide | 3.27 (s) | ↑ | $2.95x10^{-9}$ $2.10x10^{-8}$ | Oily fish |
| 16 | Glucose | 3·42 (m), 3·49 (m), 3·54 (dd), 3·74 (m), 3·84 (m), 3·91 (dd) | ↓ | $1.59x10^{-6}$ $5.89x10^{-6}$ | Sugars |
| 17 | Glycine | 3.57 (s) | ↓ | $1.71x10^{-7}$ $7.91x10^{-7}$ | |
| *18* | *N*-methyl-2-pyridine-5-carboxamide | 3·65 (d), 6·67 (d), 7·83 (dd), 8·34 (d) | ↑ | $1.78x10^{-5}$ $5.15x10^{-5}$ | Niacin (vitamin B3) |
| 19 | Glycolate | 3·95 (s) | ↓ | $3.76x10^{-5}$ | |

| | | | | | |
|---|---|---|---|---|---|
| | | | | 1.01x10$^{-4}$ | |
| 20 | 4-hydroxyhippurate | 3·95 (s), 6·97 (d), 7·76 (d) | ↑ | 9.57x10$^{-5}$<br>2.32x10$^{-4}$ | Fruits |
| 21 | Hippurate | 3·98 (d), 7·55 (t), 7·64 (t), 7·84 (d) | ↑ | 1.08x10$^{-6}$<br>4.15x10$^{-6}$ | Fruits, vegetables |
| 22 | Tartrate | 4·34(s) | ↑ | 3.71x10$^{-11}$<br>4.05x10$^{-10}$ | Grapes |
| *23* | *N*-methylnicotinate | 4·44 (s), 8·10 (t), 8·84 (d), 9·11 (s) | ↑ | 1.50x10$^{-10}$<br>1.44x10$^{-9}$ | Niacin (vitamin B3) |
| 24 | Proline betaine | 2.30 (m), 2.50 (m), 3.11 (s), 3.30 (s), 3.55 (m), 4.08 (m) | ↑ | 1.52x10$^{-9}$<br>1.15x10$^{-8}$ | Citrus |
| 25 | 4-cresyl sulfate | 2.35 (s), 7.20 (d), 7.28 (d) | ↓ | 1.89x10$^{-6}$<br>6.89x10$^{-6}$ | Gut microbial |
| 26 | Guanidinoacetate | 3.86(s) | ↑ | 4.41x10$^{-9}$<br>3.02x10$^{-8}$ | Poultry |
| 27 | Creatinine | 3.05 (s), 4.05 (s) | ↓ | 4.41x10$^{-9}$<br>3.02x10$^{-8}$ | - |

*Metabolites are ordered based on chemical shift. Peaks are listed only if they are in the range of the processed data. Multiplicity key is abbreviated as follows: s – singlet, d – doublet, t – triplet, q – quartet, dd – doublet of doublets, dq – doublet of quartets, 2D – two doublets, m – mulitplets ^Sign of association (↑ indicates higher excretion after Diet 1, ↓ indicates higher excretion after Diet 2). ’Only known dietary sources are listed. P-values are unadjusted, while Q-values are adjusted for False Discovery Rate. A  P-value and Q-value of < 0.05 as cut-offs for significance.

**Supplementary Table 11. Comparison of urinary metabolites levels between Diet 1 (NICE guidelines) and Diet 2 (Western) at 24 hours (Day 2), 48 hours (Day 3), and 72 hours (Day 4) post intervention (n=18)** Green cells indicate metabolites with higher levels in Diet 1, while orange cells indicate metabolites with higher levels in Diet 2.

| | n=18 | | |
|---|---|---|---|
| | 24 hours<br>P-value<br>Q-value | 48 hours<br>P-value<br>Q-value | 72 hours<br>P-value<br>Q-value |
| 1-Methylnicotinate | 3.80x10$^{-5}$<br>1.24x10$^{-4}$* | 1.50x10$^{-5}$<br>1.37x10$^{-4}$* | 1.50x10$^{-5}$<br>8.60x10$^{-5}$* |
| 1-Methylnicotinamide | 2.65x10$^{-1}$<br>1.64x10$^{-1}$ | 4.17x10$^{-1}$<br>3.26x10$^{-1}$ | 2.81x10$^{-3}$<br>5.30x10$^{-3}$* |
| Lysine | 1.20x10$^{-2}$<br>1.30x10$^{-2}$* | 2.65x10$^{-1}$<br>2.38x10$^{-1}$ | 9.87x10$^{-2}$<br>8.83x10$^{-2}$ |
| Alanine | 1.82x10$^{-2}$<br>1.69x10$^{-2}$* | 1.30x10$^{-1}$<br>1.30x10$^{-1}$ | 8.99x10$^{-1}$<br>5.88x10$^{-1}$ |
| Dimethylamine | 2.34x10$^{-3}$<br>3.79x10$^{-3}$* | 8.39x10$^{-4}$<br>3.02x10$^{-3}$* | 2.08x10$^{-2}$<br>3.22x10$^{-2}$* |
| Acetate | 6.40x10$^{-1}$<br>3.47x10$^{-1}$ | 2.08x10$^{-2}$<br>3.75x10$^{-2}$* | 2.65x10$^{-1}$<br>2.25x10$^{-1}$ |
| Trimethylamine-*N*-oxide | 8.00x10-6<br>5.00x10$^{-5}$* | 3.80x10$^{-5}$<br>1.72x10$^{-4}$* | 4.20x10$^{-4}$<br>1.43x10$^{-3}$* |
| *O*-Acetylcarnitine | 3.04x10$^{-1}$<br>1.80x10$^{-1}$ | 1.30x10$^{-1}$<br>1.30x10$^{-1}$ | 9.87x10$^{-2}$<br>8.83x10$^{-2}$ |
| *N*-acetyl-*S*-methyl-cysteine-sulfoxide | 1.54x10$^{-1}$<br>1.05x10$^{-1}$ | 1.58x10$^{-3}$<br>4.74x10$^{-3}$* | 2.30x10$^{-5}$<br>9.70x10$^{-5}$* |
| Glucose | 1.20x10$^{-2}$<br>1.30x10$^{-2}$* | 5.39x10$^{-2}$<br>8.08x10$^{-2}$ | 8.14x10$^{-2}$<br>8.83x10$^{-2}$ |
| 4-hydroxyhippuric acid | 1.07x10$^{-4}$<br>2.78x10$^{-4}$* | 2.34x10$^{-3}$<br>5.25x10$^{-3}$* | 5.34x10$^{-4}$<br>1.51x10$^{-3}$* |
| Rhamnitol | 9.32x10$^{-1}$<br>4.65x10$^{-1}$ | 9.87x10$^{-2}$<br>1.11x10$^{-1}$ | 2.68x10$^{-2}$<br>3.80x10$^{-2}$* |
| Carnitine | 9.66x10$^{-1}$<br>4.65x10$^{-1}$ | 5.39x10$^{-2}$<br>8.08x10$^{-2}$ | 4.01x10$^{-3}$<br>6.81x10$^{-3}$* |
| 3-methylhistidine | 6.58x10$^{-3}$<br>9.50x10$^{-3}$* | 1.93x10$^{-3}$<br>4.96x10$^{-3}$* | 4.68x10$^{-1}$<br>3.62x10$^{-1}$ |
| 1-methylhistidine | 9.87x10$^{-2}$<br>7.55x10$^{-2}$ | 6.65x10$^{-2}$<br>9.21x10$^{-1}$ | 9.87x10$^{-2}$<br>8.83x10$^{-2}$ |
| 3-aminoisobutyrate | 6.65x10$^{-2}$<br>5.77x10$^{-2}$ | 4.68x10$^{-1}$<br>3.51x10$^{-1}$ | 9.66x10$^{-1}$<br>6.08x10$^{-1}$ |
| Proline betaine | 1.50x10$^{-5}$ | 3.80x10$^{-5}$ | 8.00x10$^{-6}$ |

| | | | |
|---|---|---|---|
| | $6.60 \times 10^{-5}$* | $1.72 \times 10^{-4}$* | $6.50 \times 10^{-5}$* |
| Tartrate | $8.00 \times 10^{-6}$<br>$5.00 \times 10^{-5}$* | $8.00 \times 10^{-6}$<br>$1.37 \times 10^{-4}$* | $8.00 \times 10^{-6}$<br>$6.50 \times 10^{-5}$* |
| Lactate | $1.81 \times 10^{-1}$<br>$1.18 \times 10^{-1}$ | $4.95 \times 10^{-1}$<br>$3.56 \times 10^{-1}$ | $6.71 \times 10^{-1}$<br>$4.77 \times 10^{-1}$ |
| Citrate | $3.47 \times 10^{-1}$<br>$1.96 \times 10^{-1}$ | $4.17 \times 10^{-1}$<br>$3.26 \times 10^{-1}$ | $4.42 \times 10^{-1}$<br>$3.58 \times 10^{-1}$ |
| Formate | $3.28 \times 10^{-4}$<br>$7.11 \times 10^{-4}$* | $7.37 \times 10^{-2}$<br>$9.47 \times 10^{-2}$ | $4.32 \times 10^{-2}$<br>$5.64 \times 10^{-2}$ |
| Valine | $8.97 \times 10^{-3}$<br>$1.17 \times 10^{-2}$* | $1.39 \times 10^{-2}$<br>$2.77 \times 10^{-2}$* | $8.39 \times 10^{-4}$<br>$1.78 \times 10^{-3}$* |
| Isoleucine | $9.87 \times 10^{-2}$<br>$7.55 \times 10^{-2}$ | $3.69 \times 10^{-1}$<br>$3.16 \times 10^{-1}$ | $5.39 \times 10^{-2}$<br>$6.54 \times 10^{-2}$ |
| Leucine | $1.58 \times 10^{-3}$<br>$2.93 \times 10^{-3}$* | $1.81 \times 10^{-1}$<br>$1.72 \times 10^{-1}$ | $6.71 \times 10^{-4}$<br>$1.63 \times 10^{-3}$* |
| Tyrosine | $9.66 \times 10^{-1}$<br>$4.65 \times 10^{-1}$ | $8.65 \times 10^{-1}$<br>$5.77 \times 10^{-1}$ | $7.02 \times 10^{-1}$<br>$4.77 \times 10^{-1}$ |
| 4-hydroxyphenylacetate | $1.59 \times 10^{-2}$<br>$1.59 \times 10^{-2}$* | $8.14 \times 10^{-1}$<br>$9.77 \times 10^{-1}$ | $9.87 \times 10^{-2}$<br>$8.83 \times 10^{-2}$ |
| Succinate | $1.42 \times 10^{-1}$<br>$1.02 \times 10^{-1}$ | $7.99 \times 10^{-1}$<br>$5.53 \times 10^{-1}$ | $7.02 \times 10^{-1}$<br>$4.77 \times 10^{-1}$ |
| Hippurate | $1.45 \times 10^{-4}$<br>$5.80 \times 10^{-4}$* | $8.39 \times 10^{-4}$<br>$2.10 \times 10^{-4}$* | $1.93 \times 10^{-3}$<br>$2.32 \times 10^{-3}$* |
| Phenylacetylglutamine | $6.71 \times 10^{-1}$<br>$5.36 \times 10^{-1}$ | $3.85 \times 10^{-2}$<br>$7.70 \times 10^{-3}$* | $3.85 \times 10^{-2}$<br>$3.85 \times 10^{-2}$* |
| 4-cresyl sulfate | $2.84 \times 10^{-1}$<br>$2.84 \times 10^{-1}$ | $2.52 \times 10^{-4}$<br>$8.40 \times 10^{-5}$* | $1.29 \times 10^{-3}$<br>$1.93 \times 10^{-3}$* |
| Hip:PAG | $6.58 \times 10^{-3}$<br>$1.32 \times 10^{-2}$* | $1.45 \times 10^{-4}$<br>$8.40 \times 10^{-5}$* | $7.60 \times 10^{-5}$<br>$2.14 \times 10^{-4}$* |
| Hip:4CS | $3.85 \times 10^{-2}$<br>$5.13 \times 10^{-2}$ | $2.52 \times 10^{-4}$<br>$8.40 \times 10^{-5}$* | $2.30 \times 10^{-5}$<br>$1.37 \times 10^{-4}$* |
| PAG:4CS | 1.00<br>$6.67 \times 10^{-1}$ | $2.29 \times 10^{-1}$<br>$3.81 \times 10^{-2}$ | $1.07 \times 10^{-4}$<br>$2.14 \times 10^{-4}$* |

* Significant variables fulfilling both p-value and FDR adjusted q-value thresholds of < 0.05.
Hip, hippurate; PAG, phenylacetylglutamine; 4CS, 4-cresyl sulfate.

**Supplementary Figure**

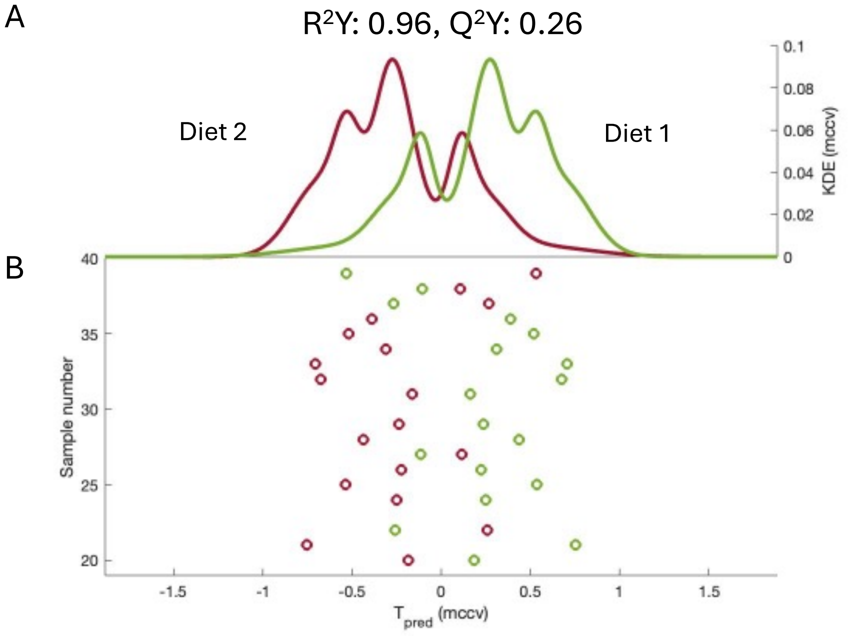


**Figure.S1 RM-MCCV-PLS-DA model comparing the global serum profiles after 72-hour adherence to Diet 1 (NICE guideline) and Diet 2 (Western) (n=18).** (A) Kernel density estimate of the predicted scores for Diet 1 (green) and Diet 2 (red), (B) Mean predicted score from MCCV for each individual. RM-MCCV-PLS-DA, Repeated-measures partial least squares discriminant analysis model using Monte Carlo cross-validation.
